# Public–Private Mix (PPM) for Tuberculosis (TB) in Urban Health Systems in Least Developed, Low Income and Lower-Middle-Income Countries and Territories – A Systematic Review

**DOI:** 10.1101/2024.05.01.24306566

**Authors:** Aishwarya Lakshmi Vidyasagaran, Noemia Teixeira de Siqueira, Sampurna Kakchapati, Thomas Falconer Hall, Baby Naznin, Jannatun Tajree, Zahidul Quayyum, Deepak Joshi, Florence Tochukwu Sibeudu, Pamela Adaobi Ogbozor, Ifeyinwa Ngozi Arize, Grishu Shrestha, Su Golder, Maisha Ahsan, Swaksar Adhikary, Prince Agwu, Helen Elsey

**Affiliations:** Research Associate, Department of Health Sciences, University of York, York, UK; Research Fellow, Department of Health Sciences, University of York, York, UK; Research Manager, HERD International, Lalitpur, Nepal; Public Health Registrar/Visiting Research Fellow, Department of Health Sciences, University of York, York, UK; Senior Research Associate, BRAC James P Grant School of Public Health, BRAC University, Dhaka, Bangladesh; Senior Research Assistant, BRAC James P Grant School of Public Health, BRAC University, Dhaka, Bangladesh; Professor, BRAC James P Grant School of Public Health, BRAC University, Dhaka, Bangladesh; Operational Research and Evaluation Manager, HERD International, Lalitpur, Nepal; Reader, Department of Nursing Science, Nnamdi Azikiwe University, Awka, Nnewi Campus, Nnewi, Nigeria; Lecturer, Department of Psychology, Enugu State University of Science and Technology, Enugu, Nigeria; lecturer, Department of Health Administration and Management, University of Nigeria, Nsukka, Enugu Campus, Nigeria; Research Officer, HERD International, Lalitpur, Nepal; Associate Professor, Department of Health Sciences, University of York, York, UK, YO10 5DD; Research Assistant, ARK Foundation, Dhaka, Bangladesh; Research Assistant, BRAC James P Grant School of Public Health, BRAC University, Dhaka, Bangladesh; Health Policy Research Group, University of Nigeria; School of Humanities, Social Sciences, and Law, University of Dundee; Professor of Global Public Health, Department of Health Sciences, University of York, UK; Hull York Medical School, UK

## Abstract

**Objective:** To assess the impact of Public-Private Mix (PPM) models for Tuberculosis (TB) diagnosis and treatment on health, process, and system outcomes within urban contexts of least developed, low Income, and lower-middle-income countries and territories (LMICs).

**Design:** Systematic review.

**Study selection:** Ten electronic databases and research repositories, covering published and grey literature were searched on 15 August 2022. All primary studies on PPM models delivering TB services in urban health sectors of eligible countries were included. There were no restrictions applied by type of outcome measurement, publication date, or language.

**Data extraction and synthesis:** Data were extracted on COVIDENCE and quality appraisals were carried out using the Mixed Methods Appraisal Tool (MMAT). Narrative synthesis was carried out by tabulating studies according to PPM model types (direct or interface), and assessing their performance on TB health, process (including cost-effectiveness) and system outcomes.

**Results:** Of the 55 included studies, covering quantitative (n=41), qualitative (n=5), and mixed-method (n=9) designs, the majority were from South-East Asia (n=36). PPM models had overall positive results on TB treatment outcomes, access and coverage, and value for money. They also promoted and improved TB health workers’ skills and service delivery. Most outcomes tended to favour interface models, albeit with considerable heterogeneity. Inconsistent implementation of NTP guidelines, uncoordinated referrals, and lack of trust among partners were identified as areas of improvement. Evidence was lacking on involvement of informal providers within PPM models.

**Conclusions:** PPM models can be effective and cost-effective for TB care in urban LMIC contexts, particularly when levels of mistrust between public and private sectors are addressed through principles of equal partnership. The evidence indicates that this may be more achievable when an interface organisation manages the partnership.

**Study registration:** PROSPERO CRD42021289509.

**Key messages:** **What is already known on this topic?**

-Although previous reviews have concluded overall improvements in Tuberculosis (TB) service outcomes with Public-Private Mix (PPM) implementation, they did not explicitly focus on urban contexts. Given the rate of urbanisation in low-and middle-income countries (LMICs) and the proliferation of PPs in urban areas, an up-to-date synthesis of the urban-specific evidence is needed for policy makers to design effective PPMs.

**What this study adds**

-Following recommended guidelines for conducting systematic reviews, we have narratively synthesised the evidence on the impact of TB-PPM models across health, process, and system outcomes within urban contexts of LMICs.

-The implemented models appear cost-effective form the societal perspective and contribute to better TB treatment outcomes, and increased access and coverage. They also consistently promote TB health workers’ skills and service delivery. Mistrust between public and private sectors can be addressed through regular communications built on principles of equal partnership.

-Although most results tend to favour models managed by interface organisations, the high heterogeneity and poor quality-scores of reporting studies must be considered.

**How this study might affect research, practice, or policy**

-This context-specific mixed-methods systematic review supports the implementation of PPM models for TB care in cities in LMICs. Providing decision-makers with evidence on the best design of PPM models is, however, less straightforward.

-Our review supports the need for more studies assessing different PPM model types, as well as clearer and more standard reporting of models and their performances.

-Very few studies mentioned the inclusion of informal providers in PPM-TB models. These providers have an important role in providing healthcare for vulnerable urban populations in the LMIC contexts. This gap must be addressed in future discussions and planning of TB-PPM models.

## Introduction

Tuberculosis (TB) remains a major global health problem despite being preventable and treatable. According to the World Health Organization (WHO), in 2022, there were 10.6 million cases and 1.6 million deaths due to TB world over (1). The impacts of COVID-19 have led to increases in TB deaths for the first time in over a decade (1). Severe disruptions in routine TB care and services in many countries have set back progress towards achieving the WHO End TB Strategy goals of reducing TB deaths and incidence (2, 3). This is to the extent that meeting the Sustainable Development Goal of ending the TB epidemic by 2030 now seems unlikely (4, 5). Many countries also face profound economic and health losses due to the additional TB burden (5).

If progress is to be made towards the global goal of eradicating TB, expanding access to TB diagnosis and treatment services is clearly a priority action (6). Between 2015 and 2020, there were an estimated 3 to 4 million ‘missing people with TB.’ This refers to the difference between cases reported in national data and estimates of TB prevalence. These ‘missing people’ are assumed to have been treated in the private sector (1). It is not surprising, therefore, that a key priority for the Global Stop TB Partnership and the WHO is to scale up public-private mix (PPM) models, with a specific focus on improving TB care and data reporting in the private health sector (6).

Nowhere is the need to partner with the private sector clearer than in cities in low-and middle-income countries (LMICs). Rapidly growing urban populations have outstripped the capacity of meagre existing public services to meet healthcare demands (7). Studies have consistently shown that city dwellers, particularly the poorest, rely on a plethora of private providers (PPs) (8, 9). This is particularly true of TB, which is fuelled by the very determinants that are prevalent in poor urban neighbourhoods, such as overcrowding, poor nutrition, and high tobacco use (10). In addition, the private sector, characterised by limited regulation and widely available over-the-counter anti-TB drugs, tends to be the most common first point of contact for people with TB (11, 12). This combination of high vulnerability to TB and easy access to PPs for treatment underlines the need for city governments and national TB programmes (NTPs) to find ways of harnessing the private sector to provide effective TB diagnosis and care in urban areas in LMICs.

PPM models are a well-recognised and recommended mechanism to address these challenges and improve TB treatment outcomes; previous reviews have concluded overall improvements in TB service outcomes, especially in resource-limited areas (13–15). However, the reviews did not explicitly focus on urban contexts, which is key to address, given the rapid urbanisation and proliferation of PPs in urban areas and the need for policy makers to understand how to design PPMs for urban contexts (16). We, therefore, aimed to describe and investigate the impact of existing PPM models for TB diagnosis and treatment on health, process, and system outcomes in urban health systems in LMICs.

## Methods

This review was developed as part of the CHORUS Research Programme Consortium which aims to develop and test ways to improve the health of the poorest urban residents and build research capacity in LMICs. This is a collaborative study developed by representatives from the CHORUS partner organisations in the UK, Nepal, Bangladesh, Ghana, and Nigeria.

Our report follows the Preferred Reporting Items for Systematic Reviews and Meta-Analysis (PRISMA) guidelines (17) (Appendix 1). The review protocol was registered on PROSPERO (CRD42021289509) (18) and published (19).

### Search strategy

We searched the following ten electronic databases and research repositories, covering published and grey literature on 15 August 2022: EMBASE, MEDLINE, Health Management Information Consortium (HMIC), Social Sciences Citation Index (SSCI), Science Citation Index (SCI), Emerging Sources Citation Index (ESCI), CENTRAL, Database of Disability and Inclusion Information Resources, WHO Library Database (WHOLIS) and 3ie. The search strategy consisted of two main facets, namely, PPM models and the countries or regions of interest (see Appendix 2 for details). No language or date restrictions were applied. In addition, reference lists of relevant systematic reviews were checked to identify any additional research, alongside screening of references and forward citations of included studies. Retrieved records were de-duplicated in EndNote and uploaded to COVIDENCE (www.covidence.org) for further evaluation.

### Inclusion criteria and study selection

We included all primary studies examining PPM models delivering TB services in urban health sectors of the 2021 World Bank-defined (20) least-developed, low-income, and lower-middle-income countries and territories. PPM models were defined as long-term (not one-off events), formal or informal arrangements between the public and private sectors, while the term ‘urban’ referred to all semi-urban, peri-urban, suburban, and urban slum and non-slum areas in eligible countries. Studies that included urban and rural areas were eligible only if urban-specific results were separately reported (see Appendix 3 for detailed eligibility criteria). A total of 17 reviewers were involved in the study selection process. Sets of two reviewers independently screened the studies, first by title and abstract, then by full texts; discrepancies were resolved through discussion with a third reviewer.

### Data extraction and synthesis

Seventeen reviewers were involved in data extraction and used a pre-piloted template specifically developed for this review and uploaded to COVIDENCE. All extractions were completed by a single reviewer and checked by a second reviewer. Quality appraisals were carried out using the Mixed Methods Appraisal Tool (MMAT) (21), covering mixed-methods, qualitative and quantitative study designs. Extraction items included publication details, PPM partners, their roles as previously defined (22) (stewardship/ support/ service provision/ monitoring/ financing), study characteristics and sample size, TB interventions provided by the PPM, reported outcomes related to health (TB treatment outcomes), process (indicators of access, coverage, utilisation, cost, etc.) and WHO-defined (23) system building blocks (service delivery, health workforce, information, equipment, financing, governance). The authors of the included studies were not contacted for any additional data. During the extraction and coding of the six system building blocks, the team found findings pertaining to the attitudes and behaviours of those involved in the PPM and the social and organisational context. Hence, two additional themes were added.

Narrative synthesis was carried out by tabulating and summarising studies according to PPM models (i.e., whether the partnership between the public and private sectors was direct or managed through an interface agency), and reported in line with the synthesis without meta-analysis (SWiM) guidelines (Appendix 4) (24). We also assessed the performance of different PPM models on TB health, process (including cost-effectiveness) and system outcomes.

For health outcomes, we used the percentages and confidence intervals (CIs) to build forest plots (without meta-analysis) grouped by PPM models; CIs not reported in the studies were calculated using standard deviations and sample sizes. Quantitative results relating to process and system outcomes were tabulated in similar groups and summarised. For studies reporting cost and cost-effectiveness, we first summarised the study perspectives (i.e., patient, provider, public sector, societal) and outcomes (i.e., treatment success, disability-adjusted life years (DALYs) averted) reported. Details on cost calculations and reporting can be found in Appendix 5. Next, we plotted the results in a cost-effectiveness diagram.

All extracted qualitative findings were coded and grouped under the qualitative themes using Nvivo 1.7 (25). An inductive reasoning approach was adopted, with three reviewers independently coding the qualitative findings. Disagreements or discrepancies were resolved by discussion. The synthesised qualitative findings were combined with quantitative findings and reported against each building block. Finally, we conducted results-based convergent synthesis to explore how the PPP models affect their outcomes.

### Amendments to the protocol

Our original review covered all health conditions (19). However, given the large number of studies on TB control and the comparability of outcomes reported, we amended our protocol (updated on PROSPERO) to carry out additional synthesis specifically on those papers reporting urban TB PPM models. The screening criteria originally excluded tertiary healthcare settings but was amended to remove this clause after piloting for 25 studies. Finally, we did not use RE-AIM (26) for qualitative analysis as originally planned, as the framework did not adequately capture the outcomes reported in the included studies.

### Patient and public involvement

No patients or members of the public were involved in the conception, development, or analysis of the review. No patients were asked to advise on the interpretation or writing up of results.

## Results

Our searches identified 7,346 records, from which 55 eligible studies on TB (reported in 63 publications) were included (27–89) (Figure 1). For eight studies (27, 28, 33, 34, 48, 49, 57, 58, 60, 61, 70, 71, 84, 85, 87, 88), the research was reported across two publications each, which were merged and extracted as one study (Table 1 and Appendix 6). Most studies were from the WHO South-East Asia region (n=36, 65.4%) (30, 33, 36, 37, 40–42, 45, 47, 48, 50, 51, 54–56, 59, 60, 62, 64–67, 69, 74–76, 78–84, 86, 87, 89), followed by Eastern Mediterranean (n=7, 12.7%) (32, 43, 44, 46, 72, 73, 77), Western Pacific (n=6, 10.9%) (27, 31, 35, 52, 57, 70), African (n=4, 7.3%) (38, 39, 53, 68), and American (n=1, 1.8%) (63) regions. One study reported models from across three different countries and regions (29).

**Figure 1.**
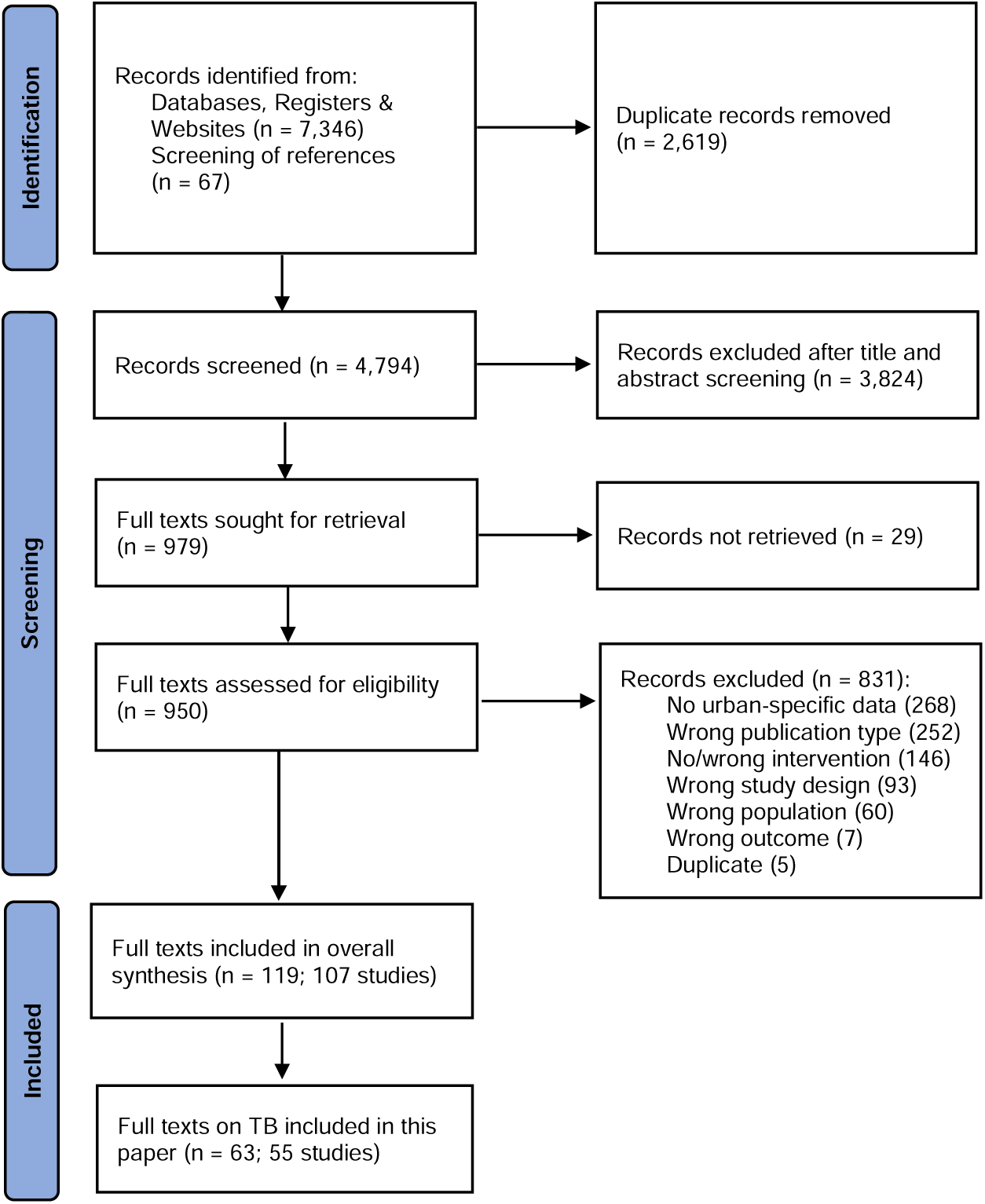
– PRISMA flow diagram

**Table 1.**
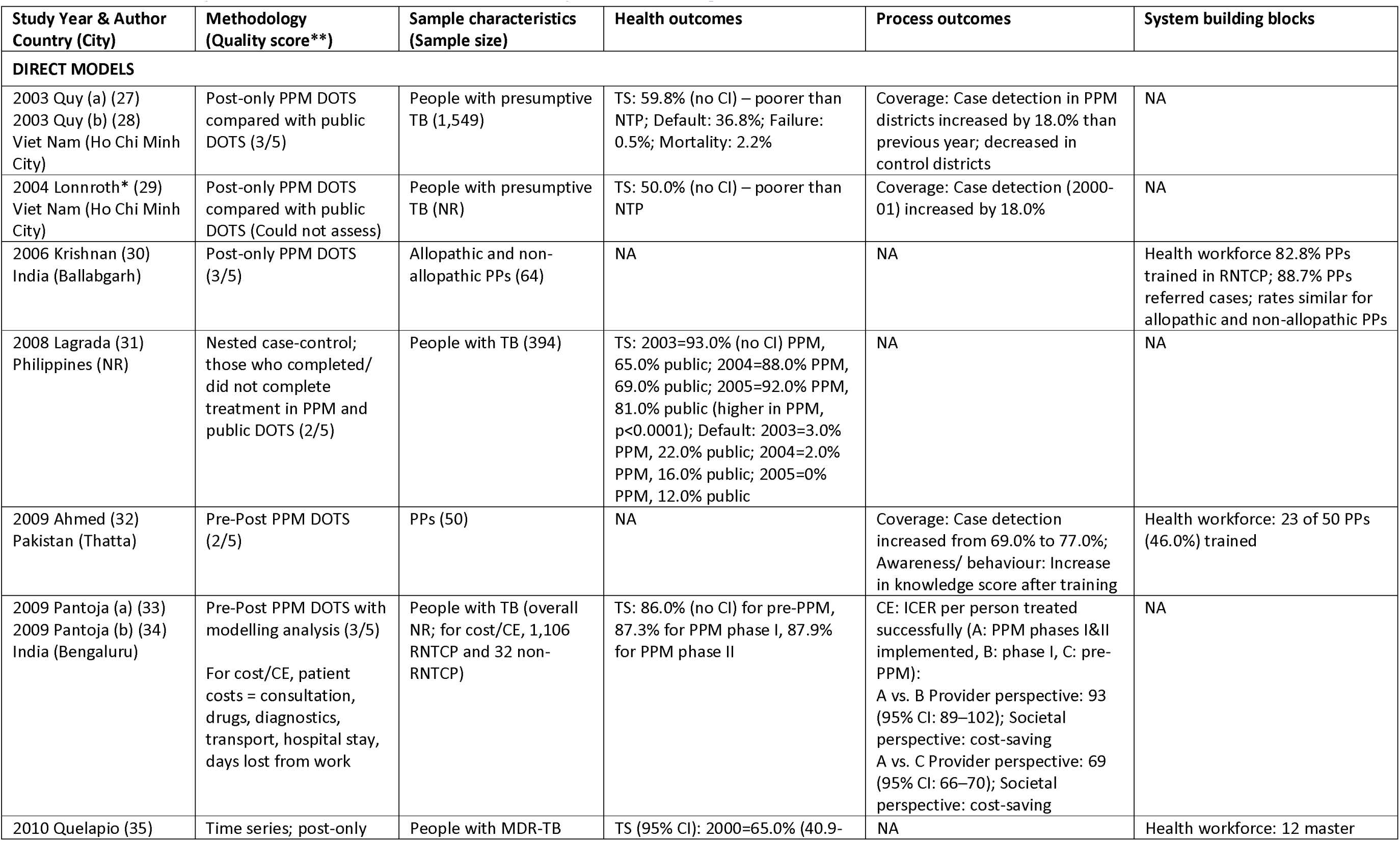
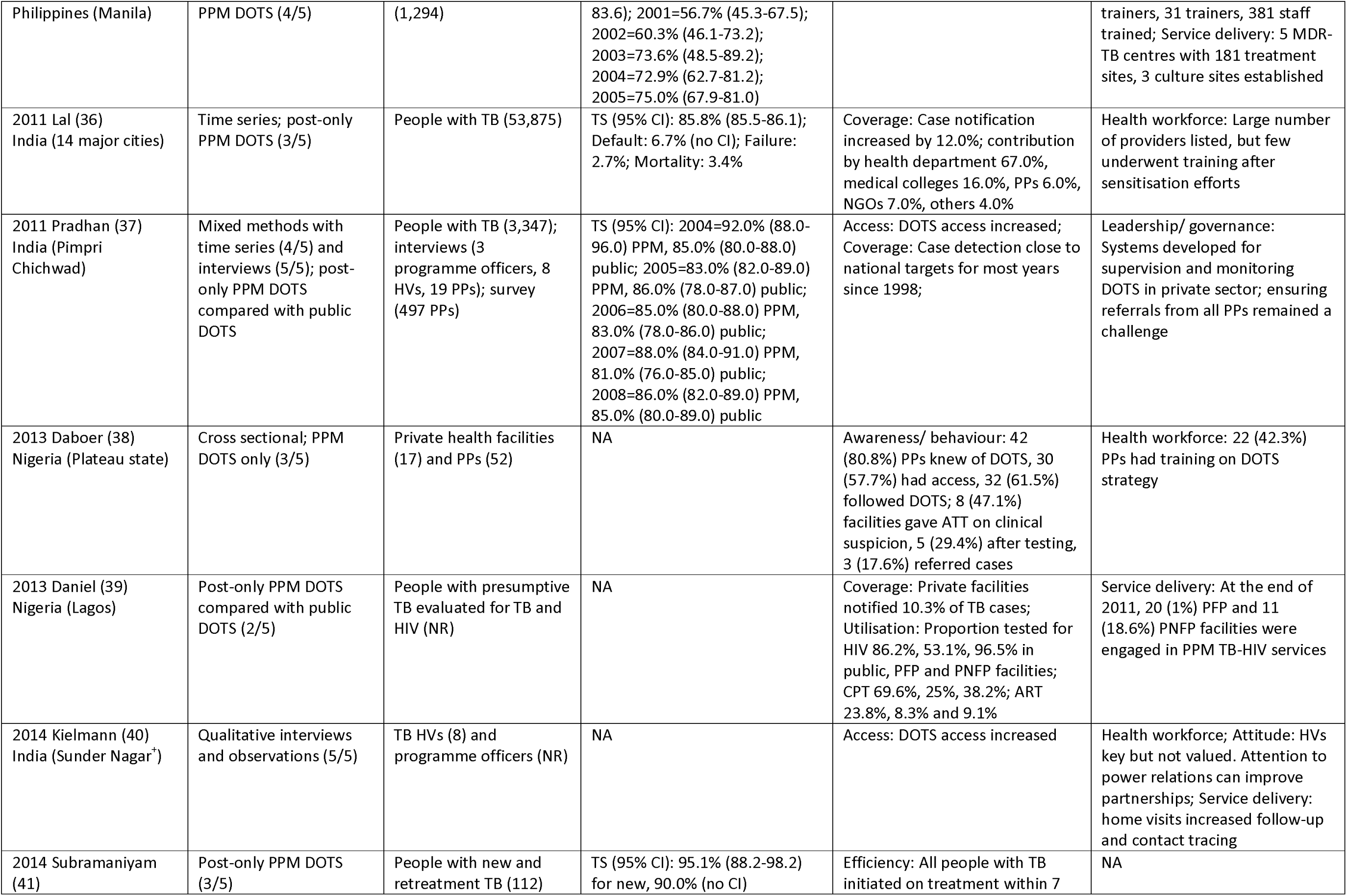
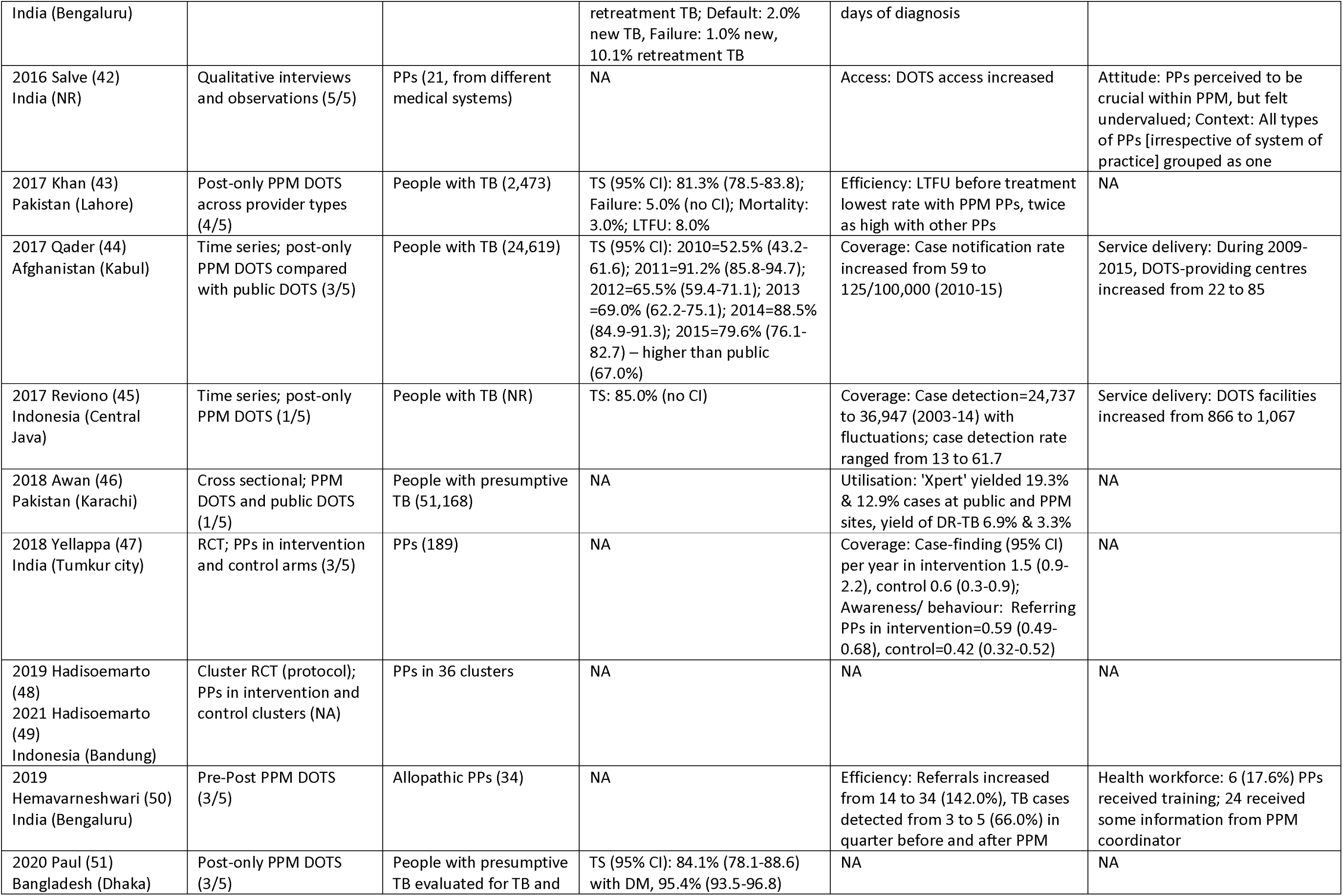
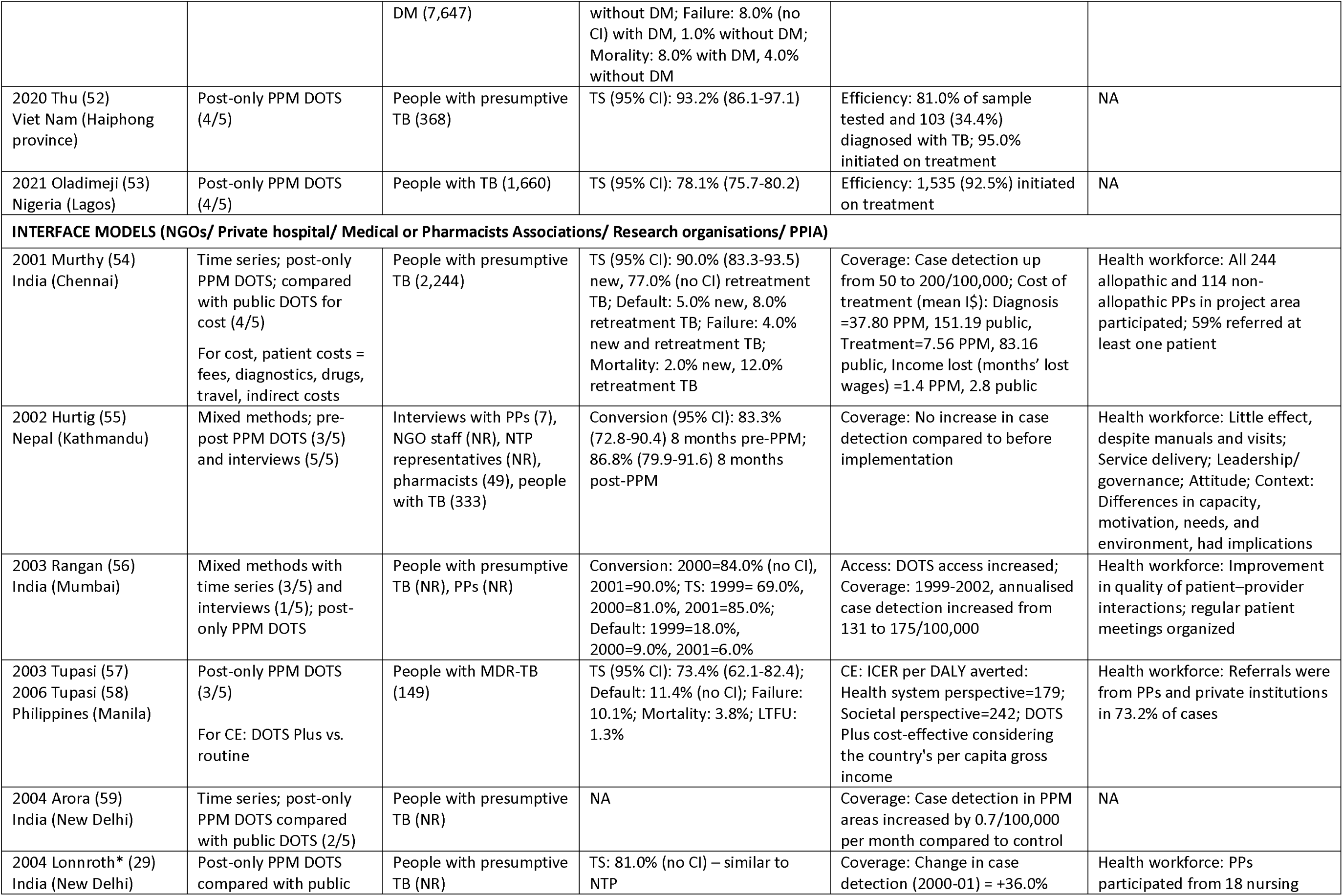
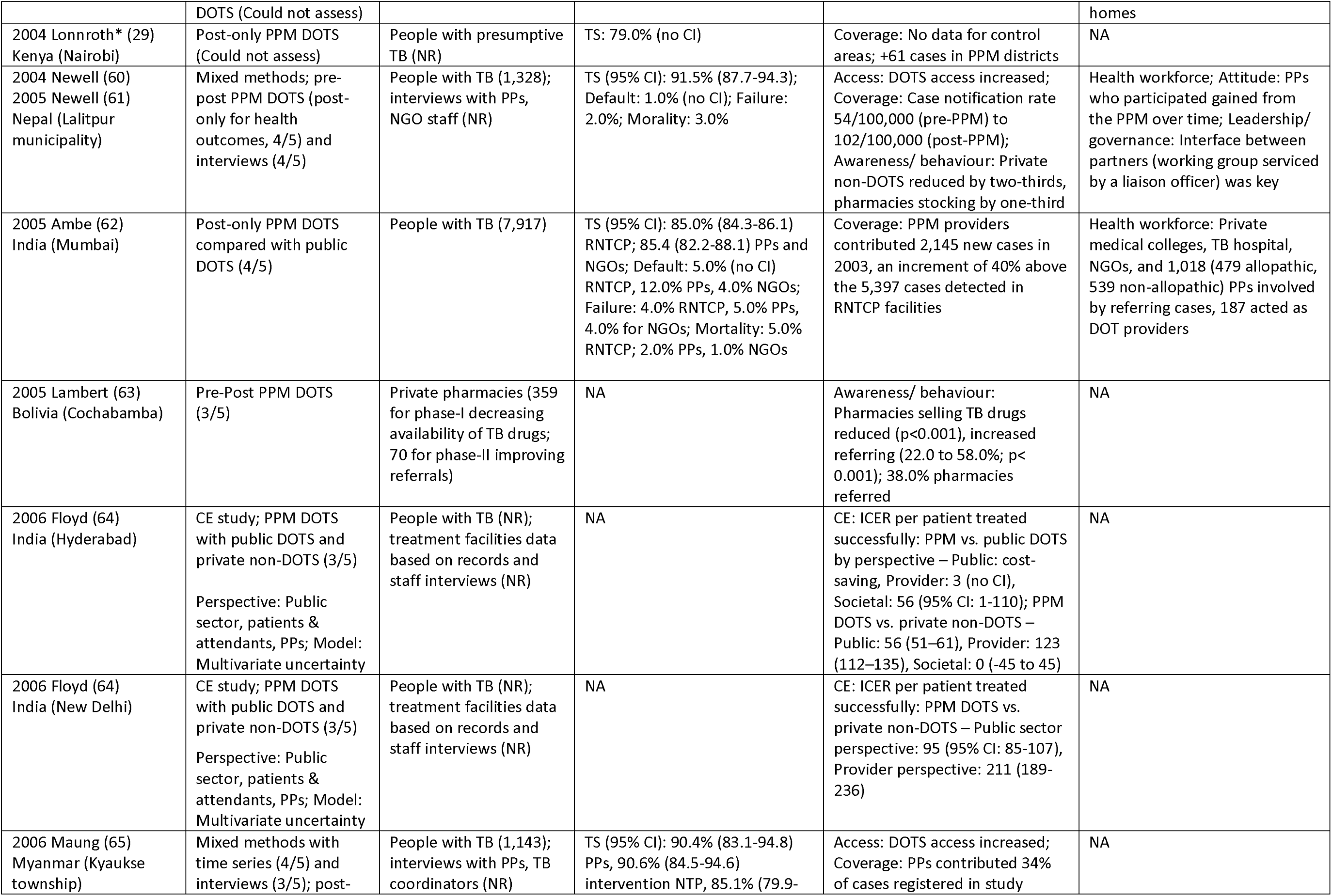
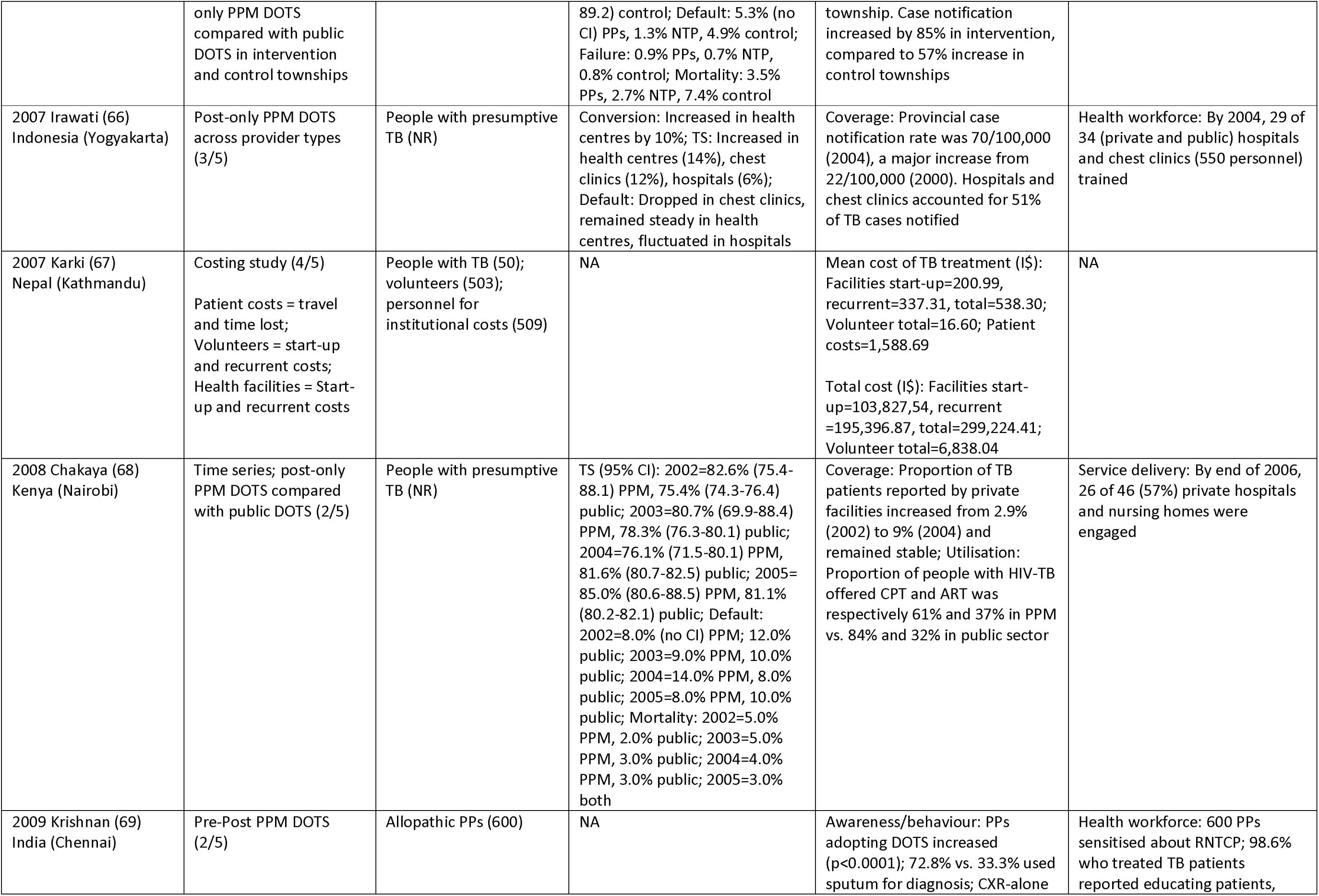
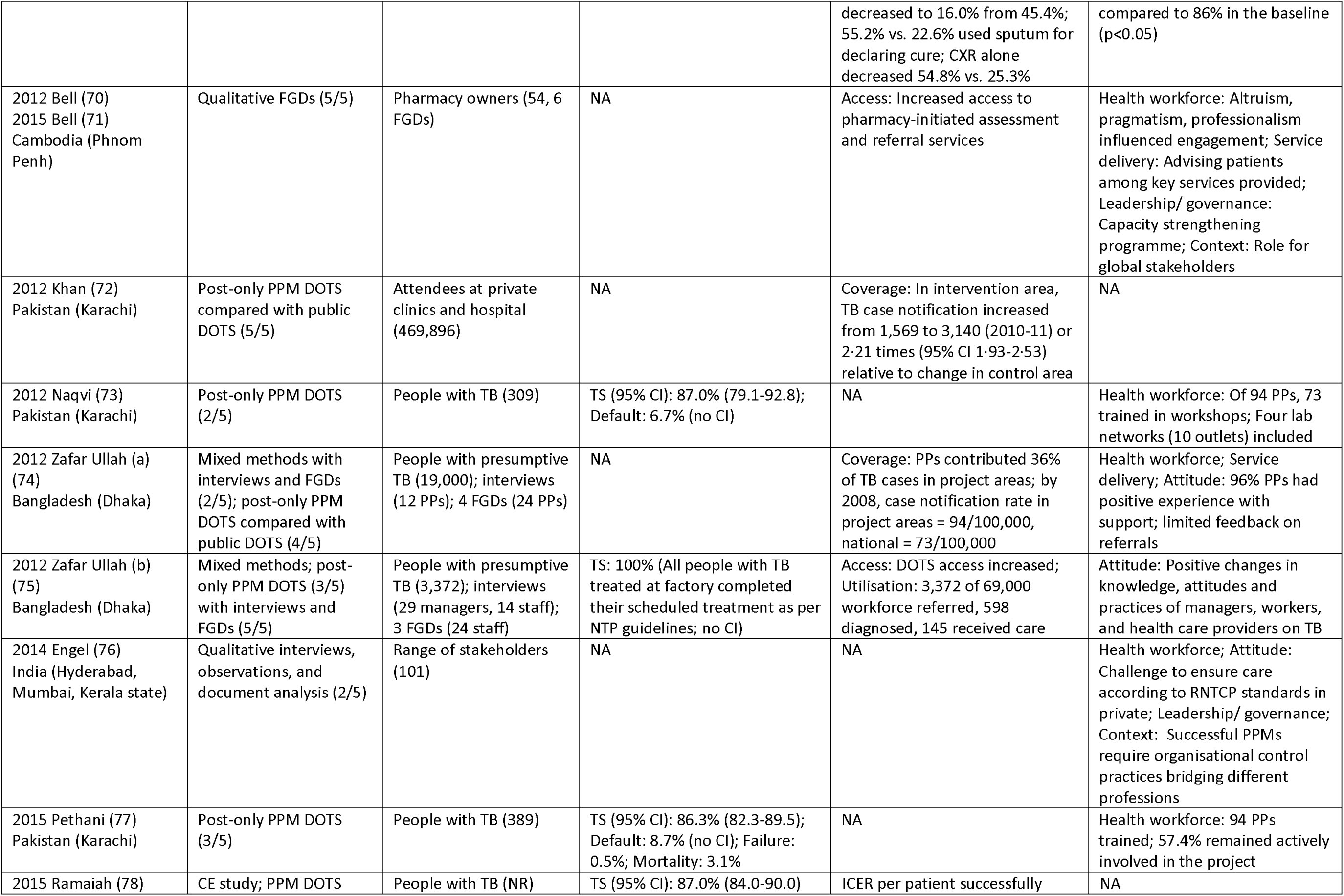
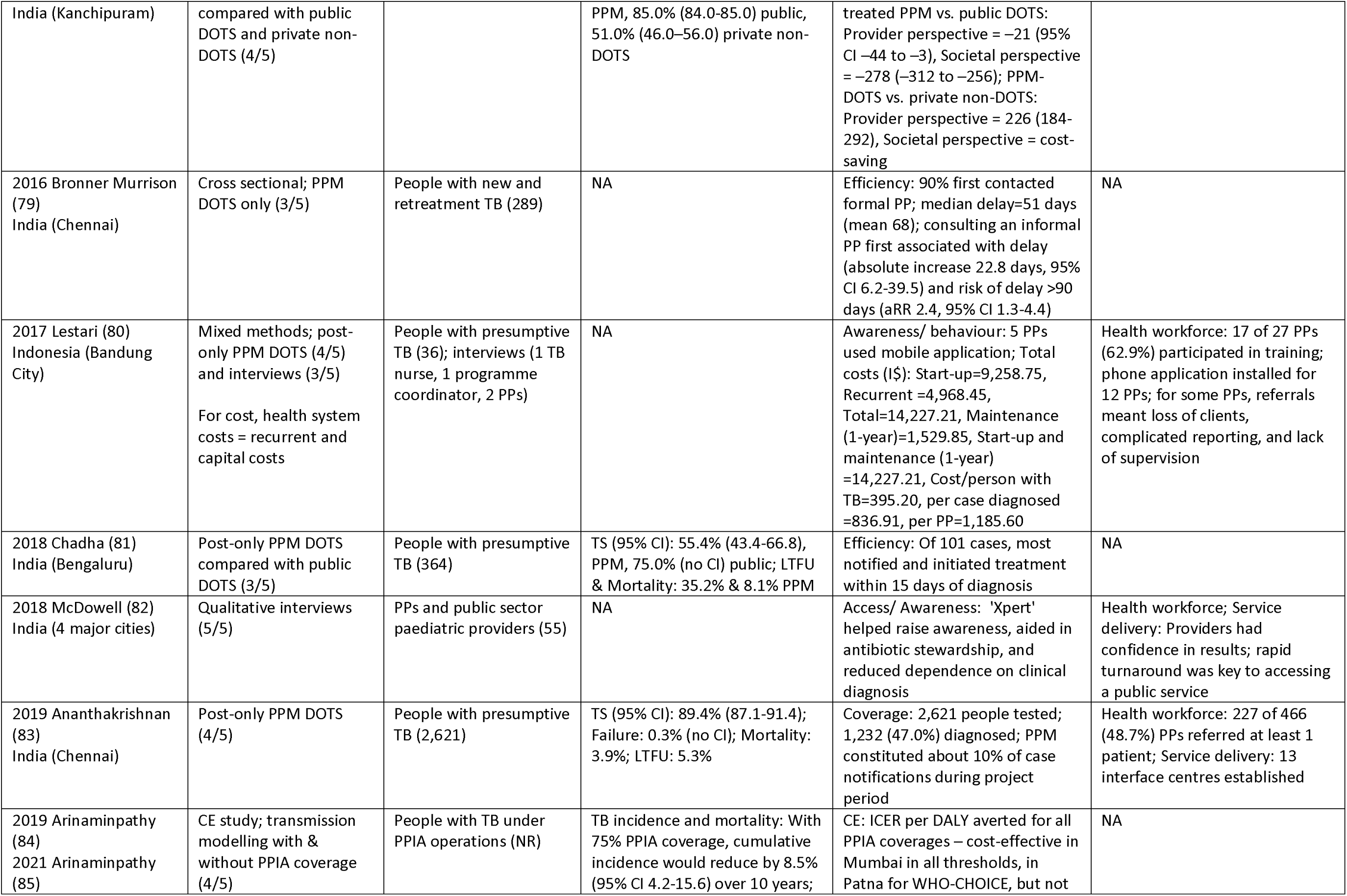
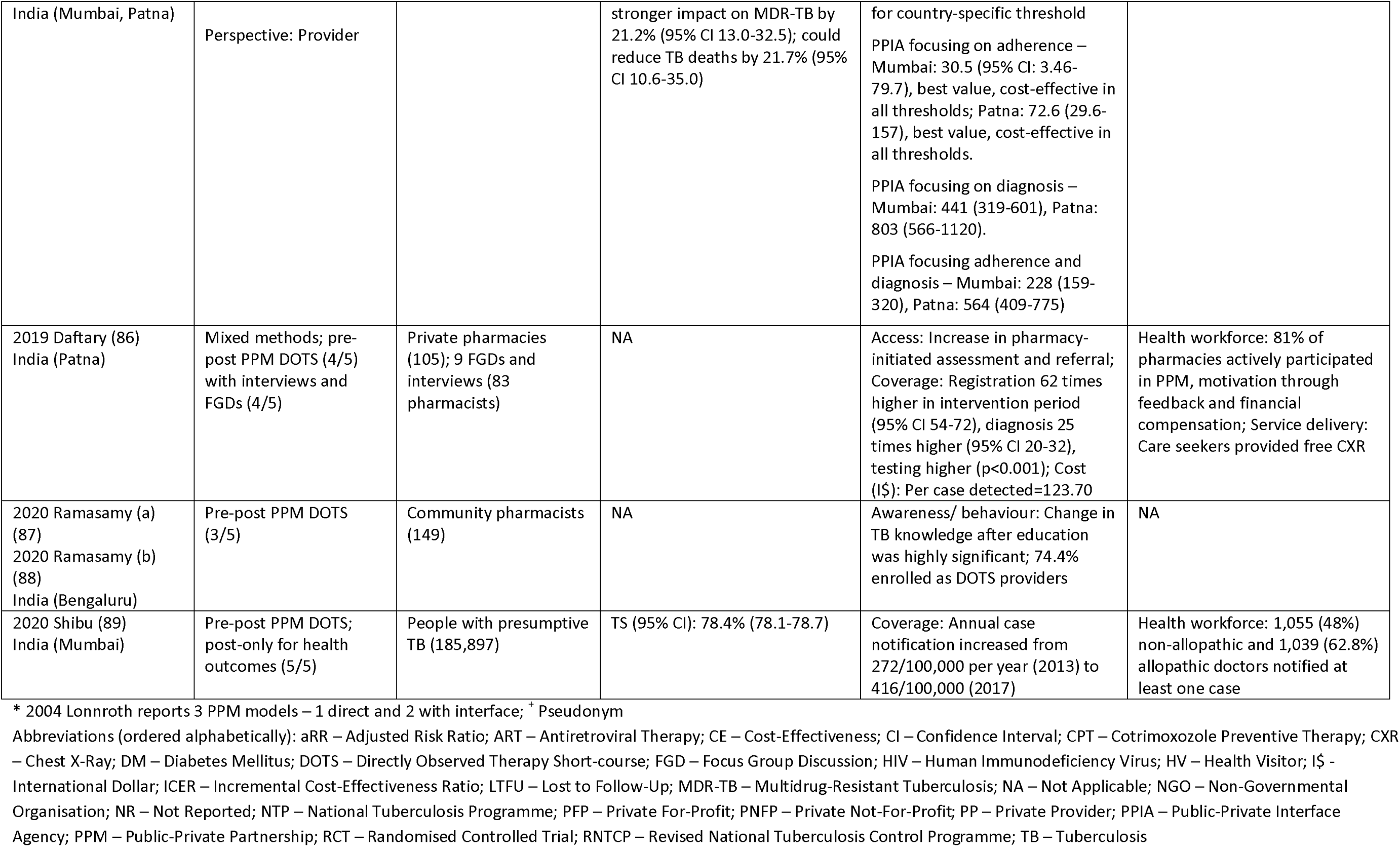
– Summary of results (Health, Process, and System building blocks)

We included quantitative (RCT protocol 1, RCT 1, non-randomised intervention studies with comparison 9, without comparison 19, observational 4, cost & cost-effectiveness 7), qualitative (n=5) and mixed-methods (n=9) studies. Few studies (mostly qualitative) met all the design-specific quality appraisal criteria (n=6, 10.9%), while scores for the remaining studies were distributed as follows: scores 4/5 = 9 (16.4%) studies, 3/5 = 19 (34.5%) studies, 2/5 = 8 (14.5%) studies, and 1/5 = 2 (3.6%) studies (Table 1). In the mixed-methods studies, we found no clear integration between the different study components, but the majority scored 3/5 or greater in the separate qualitative and quantitative parts. Finally, quality scores could not be assessed for one study protocol (48) and one study, which did not provide the necessary methodological details (29) (Table 1).

### Characteristics of PPM models included in the review

We found two types of PPM models reported (Table 1 and Appendix 6) – those in which the public sector TB programme formed a direct partnership with the private sector (Direct models, n=24), and those in which there was an interface (Non-governmental Organisations (NGOs)/ Private hospital/ Medical or Pharmacists Associations/ Research organisations/ Public-Private Interface Agency (PPIA)) between the public and private sector partners (Interface models, n=36). In both model types, the roles of stewardship, monitoring, and support (as defined by Tabrizi et al., (22)) were mainly provided by the public sector, although support was also provided by the private sector partner more often in the interface compared to the direct models. Information regarding financing was not reported in many papers, but when it was reported, the public sector partner took on this role more often. Both sets of partners provided TB services (Appendix 6). Regarding the interventions provided, while some models focused on increasing case detection, the majority also aimed to decentralise TB treatment. A mix of outcomes (health/process/system) was reported in most studies, while some only reported results relating to one outcome category. For the RCT protocol that was included (48, 49), only details of the PPM model were extracted.

### Health outcomes

Health outcomes for urban-specific populations were reported in 31 studies, comprising treatment success (n=29), unfavourable treatment outcomes such as default, failure, mortality (n=19), sputum conversion (n=3), and TB incidence (n=1).

Of the 29 studies reporting treatment success (27, 29, 31, 33, 35–37, 41, 43–45, 51–54, 56, 57, 60, 62, 65, 66, 68, 73, 75, 77, 78, 81, 83, 89), 17 (60.7%) reported rates >=85%. Figure 2 plots the estimates from 21/28 studies (for which 95% CIs were reported or could be estimated), according to PPM model type. The interface models achieved a treatment success rate close to 90% more consistently than the direct models, although given the heterogeneity of the studies, it was not possible to draw conclusions. Ten studies compared favourable and/ or unfavourable treatment outcomes (treatment success, default, failure, mortality) for PPM Directly Observed Treatment Short course (DOTS) provision with public DOTS/private non-DOTS during the study period (27, 29, 31, 37, 44, 62, 65, 68, 78, 81). The majority (8/10) reported comparable or better outcomes with the implementation of PPM models (Table 1 and Appendix 7). Regarding sputum conversion, all three studies reported increases after PPM implementation (55, 56, 66). One modelling study estimated that with 75% PPM with interface coverage, the cumulative incidence of TB would reduce by 8.5% (95% CI 4.2-15.6) over 10 years (84).

**Figure 2.**
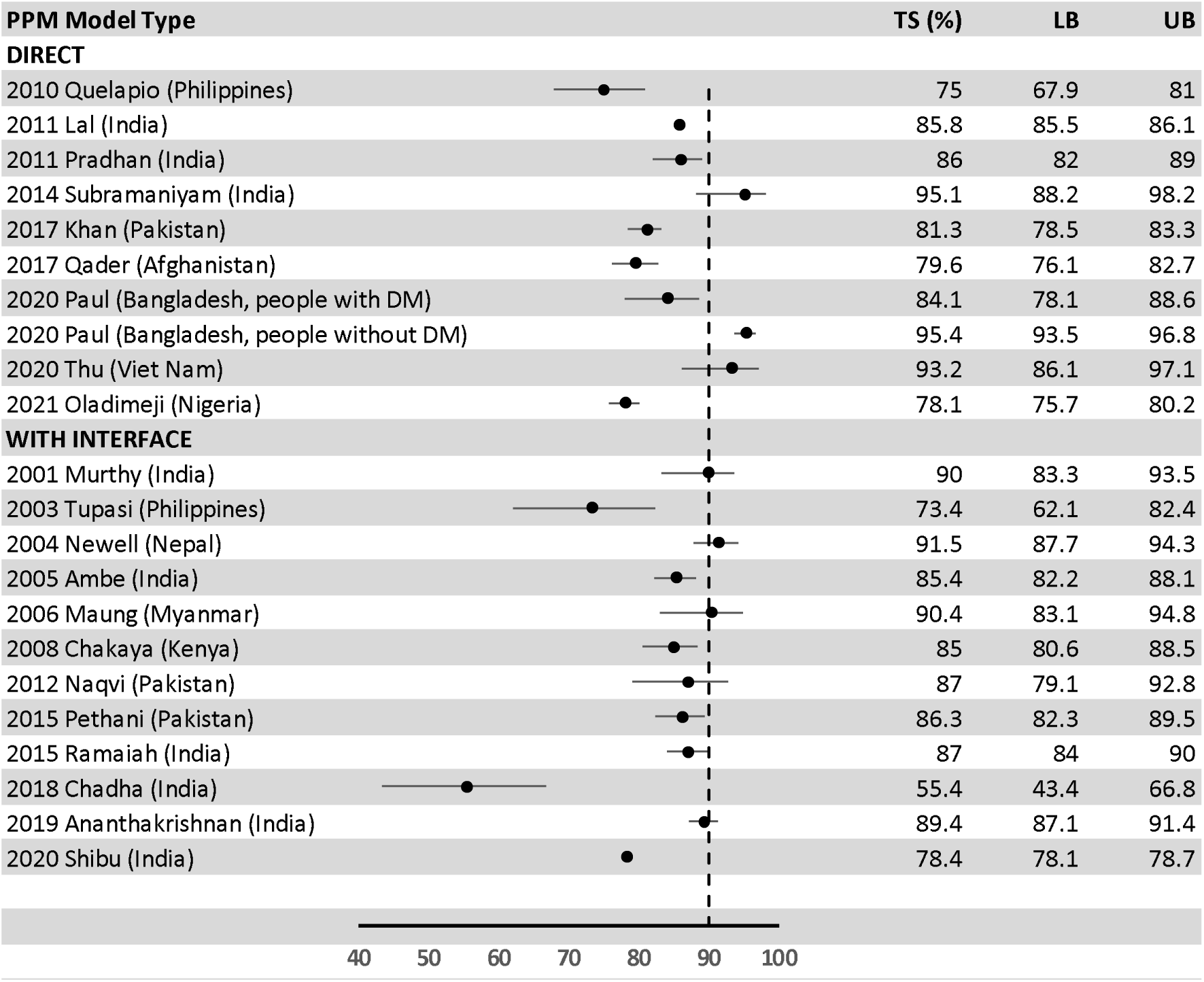
– Forest plot (without meta-analysis) of studies reporting TB treatment success according to PPM model type.

### Process outcomes

Process outcomes were reported in a total of 45 studies, including access (n=10), coverage (case detection/ notification) (n=23), utilisation (n=4), awareness/ behaviour change (n=9), and improved efficiency (n=7). Further, we found studies that reported cost (n=4), cost-effectiveness (n=4), and both cost and cost-effectiveness (n=1).

Results showed an increase in access to TB diagnostic services (including GeneXpert testing) (70, 82, 86) and DOTS (37, 40, 42, 56, 60, 65, 75) with PPM implementation. Almost all studies reporting coverage (in both direct and interface models) reported an increase in case detection/ notification (up to 40% (62)) with PPM implementation, or they found higher rates in PPM compared to control settings (47, 59, 65, 72). In only one study, no increase in case detection was found (55), while in another, it was only reported as being close to national targets (37). On the other hand, we found less favourable results relating to utilisation of TB diagnostic and treatment services within the context of PPMs. TB testing based on GeneXpert (46), as well as TB-HIV testing and treatment (39, 68), appeared to be poorer in PPM compared to public settings. In one PPM model providing workplace DOTS, only 24.2% of those diagnosed with TB undertook treatment at the workplace (75).

Awareness and behaviour change among PPs (including pharmacies) were mainly reported in interface models and positively influenced by PPMs. These included greater use of sputum testing, decreased reliance on chest X-rays, and adoption of DOTS among PPs (60, 69, 80, 82, 87). In private pharmacies, reduction in stocking and selling of anti-TB drugs, and greater referral to NTP services were noted (60, 63). Among the direct models, one cross-sectional study found that a considerable number of PPs still lacked access to DOTS guidelines (42.3%) and continued to treat TB on clinical suspicion (47.1%) (38). Nonetheless, increase in TB knowledge (32) and greater referrals to NTP services were also found among PPs after implementation of two direct PPM models (47).

Regarding the outcome ‘improved efficiency’, high TB treatment initiation was consistently reported across both model types (41, 52, 53, 81). Studies also reported lower loss to follow-up before treatment initiation among PPM PPs compared to non-PPM PPs (43). In one interface model which included both formal and informal PPs, consulting an informal PP first was associated with delay in diagnosis (absolute increase 22.8 days, 95% CI 6.2-39.5) and increased risk of long delays (aRR 2.4, 95% CI 1.3-4.4) (79).

### Cost and cost-effectiveness studies

Among the studies reporting cost and cost-effectiveness, only one (reporting both outcomes) was based on a direct model (34), while all remaining studies evaluated interface models (54, 58, 64, 67, 78, 80, 85, 86). The cost studies looked at out-of-pocket expenses, time and income loss incurred by patients (n=2), as well as costs incurred by the PPM providers to implement the intervention (n=3). Most cost-effectiveness studies (n=4) adopted the societal perspective.

One costing study in Hyderabad, India (interface model), found lower out-of-pocket expenditures (e.g., fees, transport, diagnostic investigations, medications) and income loss (months’ lost wages) for patients treated under PPM-DOTS compared to public-sector DOTS (treatment costs: I$ 7.56 vs I$ 83.16; months’ lost wages: 1.4 vs 2.8) (54). Another before and after costing study from India found lower out-of-pocket expenditures for patients after implementing a direct PPM model to aiming ensure good quality TB diagnosis and treatment (I$ 130.03 vs I$ 897.80) (34). From the health system perspective (all interface models), the total cost (start-up plus one-year recurrent costs) of implementing PPM-TB varied substantially from I$ 14,227 for an intervention aiming to increase TB case detection, diagnosis, and treatment in Bandung City, Indonesia (80) to I$ 299,224 for an intervention aiming to improve the quality and coverage of TB diagnosis and treatment in Latipur City, Nepal (67). Costs to PPM provides per TB case detected was calculated to be I$ 424.42 in Patna, India (86) and I$ 836.91 in Bandung, Indonesia (80) (Appendix 8).

Amongst the cost-effectiveness studies, one reported the incremental cost-effectiveness ratio (ICER) per Disability Adjusted Life Year (DALY) averted (85), one the cost per DALY gained (58), and all others ICER per patient successfully treated (34, 64, 78). The ICER varied according to the intervention, perspective (e.g., societal, health system, patient), cost-effectiveness threshold (i.e., WHOC-CHOICE based on the Gross National Income and country-specific threshold), and comparator, falling in different quadrants of the cost-effectiveness diagram (Figure 3). Two studies with the interface model indicated that compared to separate public and private-sector DOTS, PPM-DOTS was cost-saving (more effective and less expensive) from a societal perspective (64, 78). Another cost-saving intervention from a societal standpoint was the scale-up and intensification of a direct PPM in 14 large cities in India (34). Using the WHO-CHOICE threshold, the DOTS-Plus project targeting MDR-TB was also cost-effective from a societal perspective in the Philippines (58). From the provider perspective, an interface model aiming to provide high-quality TB diagnostic tests and maximise treatment completion, presented mixed results in India. The intervention was cost-effective for all coverages in both Patna and Mumbai when adopting the WHO-CHOICE threshold. However, when adopting a country-specific threshold, the intervention was cost-effective in Patna only when focused on improving treatment outcomes (85) (Appendix 8 and Figure 3).

**Figure 3.**
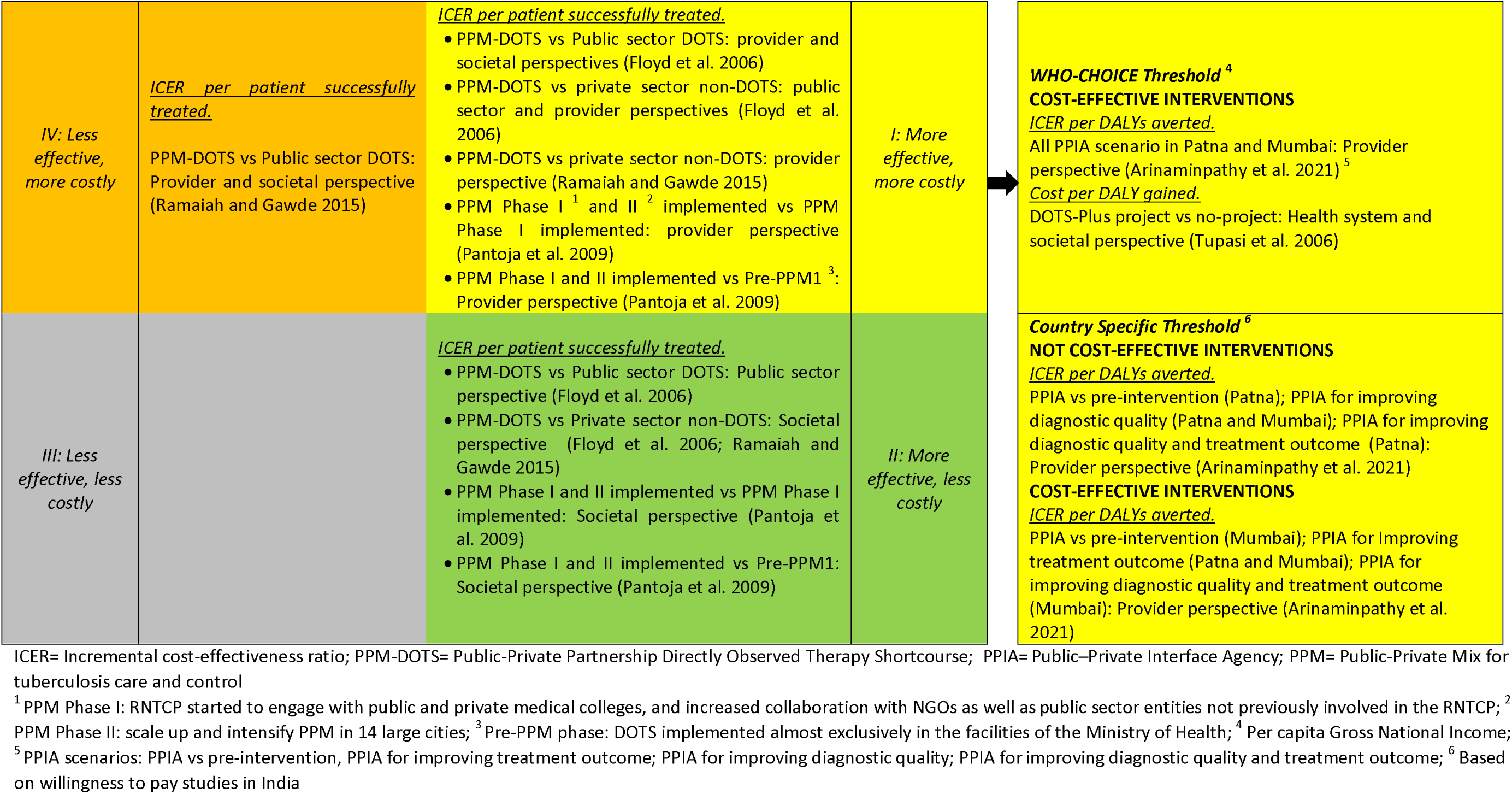
– Cost-effectiveness plan for PPM TB

### System outcomes

Results pertaining to system outcomes covered health workforce (n=26), service delivery (n=12), leadership and governance (n=5), as well as the two additional qualitative themes, namely attitudes and behaviours within the system (n=7), and context (n=4).

### Health Workforce

Several studies (both quantitative and qualitative) indicated positive outcomes in improving the knowledge and skills of the TB health workforce, and their active engagement (30, 32, 35, 66). Among the quantitative studies, one study reported using a ‘training of trainers’ approach to improve sustainability and follow-on training for the health workforce (35). The proportion of PPs receiving training ranged from 17.6% (50) to 82.8% (30) in direct models and 62.9% (80) to 100% (54) in interface models.

Findings from the qualitative studies (all but one based on interface models) presented insights from PPs on the value of the training in increasing their confidence to counsel and motivate patients to take up their referrals, also ensuring their client flow and status in society (70). Public and private sector providers both had concerns about the TB treatment knowledge and quality of care that the other sector provided. Even after training and dissemination of guidelines, some studies (55, 76, 82) found it a challenge to ensure care according to the government TB programme standards: “The government has a programme. The private practitioners have patients” (TB consultant quoted in 2014 Engel, pg. 922 (76)). Yet, training was identified as one part of the process of building trust between the two sectors (60, 74), in addition to being reassured that they would not lose clients through mechanisms such as the public sector issuing patients with back-referral letters (42, 76).

Motivation of PPs was another recurring theme within the health workforce domain. Several mechanisms which motivated PPs were identified including altruism and religion (70), improving public health (70, 86), financial compensation and feedback (86), professionalism (80), and increased legitimacy and standing in the community (40, 60). Standing in the community was particularly enhanced when the increased knowledge and improved practice of the private providers led to treatment success:

“When patients get better, they are thankful. They have trust in our pharmacies and let their friends and relatives know. So that helps build our customer base. That’s one benefit for our business.” (Focus group with pharmacists in Cambodia, 2012 Bell, pg. 1089 (70))

### Service Delivery

All relevant studies (covering study designs and both model types) reported increases in the number of DOTS-providing centres following PPM implementation (35, 39, 44, 45, 68, 83). The inclusion of a variety of private and NGO providers within the models allowed adaptations to the standard public TB services offered. For example, in PPMs where NGOs were involved, home visits were also included to increase follow-up and conduct contact tracing thus strengthening service provision (40), while another study found more limited engagement of private facilities in TB-HIV services (39). The way services were delivered also changed with increased emphasis on counselling and advising patients to seek diagnosis and complete their treatment. This was particularly identified by PPM pharmacy owners as a key part of the services provided, although some felt that community perceptions of pharmacies as only providers of drugs undermined this role (70).

Where a PPM provided a new service, it ensured buy-in from PPs, for example in 2018 McDowell (82) and 2019 Daftary (86), where the inclusion of GeneXpert diagnostics and free chest x-rays respectively, provided care seekers with a tangible service. However, when there were gaps in vital services provided by the public sector, such as microscopy, the coordinated provision of services within the PPM would breakdown (55). While referral systems were a key mechanism for improving service delivery, several studies identified challenges such as limited feedback from the public to the private system (55, 74), or poor relationships between the private and public providers, undermining referrals to public services when needed (40). Only one study found that PPs were concerned about providing TB services due to fears of infection control (70).

### Leadership and Governance

The leadership and governance findings were multifaceted and complex. Understanding clear roles and responsibilities of partners was essential, noting that individual roles could change and develop throughout the partnership. Joint working and communication were identified as important factors for TB PPMs, and time and trust required for this to happen effectively. However, “deep rooted tensions, rivalry and suspicion”, (76) particularly between the public and private sector, have been a feature of PPMs. These factors, along with a lack of confidence in partners’ capabilities, have resulted in a requirement to “build relations [and] trust” (55) through “sustained interactions” (37), based on clear commitments to the NTP and championing of the PPM. Building relations was fundamental given that PPM governance often required “changes in standards” (76) to promote and maintain the quality of services (including laboratory diagnosis, care, and drugs), necessitating control mechanisms in an environment of little previous trust or confidence in other stakeholders within the partnership. PPM stakeholders also required sufficient capacity (including the NTP) to contribute meaningfully and sustainably. This capacity could be strengthened through progressive, flexible learning programmes such as “Cambodia’s learning-by-doing philosophy, its readiness to review and adapt policy” (71) or giving partners the opportunity to gain additional experience in management that ultimately were “relevant not only to TB control but other public health issues” (71).

Where leadership and governance were housed within NGOs or other interface organisations, the qualitative findings reported that this feature was crucial to success: “A key component of the PPP was the provision of an interface between the various partners. In our case this interface was a working group serviced by a liaison officer. The working group was a forum for potential partners to meet and plan the PPP. The commitment of the liaison officer proved key to the successful implementation of the PPP.” (2005 Newell pg. 1014 (61))

Qualitative participants in one study in India (76) explored differences between direct government-led models and an interface model led by a private hospital. They found that where government leadership was seen as too strong with rigid reporting systems and supervision, and little respect for PP’s decisions, the PPM models were problematic. Conversely, interface organisations, often under the leadership of a TB ‘champion’ (61, 76) ensured open communication and respect between public and private sectors, and were more effective.

### Attitudes and behaviours within the system

Qualitative findings emphasised tensions in attitudes between public and private sectors. Where studies reported the evolution of PPMs over time, there were interesting reflections on changing nature of these attitudes, moving from distrust and suspicion particularly in relation to quality and motivation, to a greater understanding and respect (55, 60). However, building relationships took time, and this was identified as a significant barrier when PPMs followed the direct model and leadership and coordination fell to the NTP: “The NTP lacks the time, and to a lesser extent the credibility, to do this, and it will therefore usually be necessary to find someone else to act.” (2002 Hurtig, Pg. 84 (55))

Medical hierarchies were also identified as undermining the effectiveness of PPM models, with TB healthcare workers and volunteers working directly with patients and household contacts expressing their frustration at the lack of respect, calling for a shift toward recognising their pivotal role and addressing power dynamics (40, 42). In one study, NGO health workers of the same professional level as NTP supervisors reported being treated as inferior due to their non-public sector position, and this was despite an interface model where the NGO coordinated the PPM (76). Positive monitoring and support mechanisms were also emphasised for PPM success and scale-up (74, 75). Where all credit for improvements in programme outcomes was seen as being taken solely by the government this undermined the smooth running of the programme (42).

### Context

The private sector demonstrated considerable heterogeneity, including informal providers, non-allopathic professionals, and a highly accessible, developed, and regulated pharmacy network. However, PPM policies tended to “[categorise] all types of private practitioners [irrespective of system of practice] under one broad group”, seeing them “through the same lens of financial incentive” (2016 Salve, Pg. 631 (42)). While different categories of providers had different motivators for contributing to a PPM, they were not solely financial. Further, given that “unqualified practitioners predominate in poorer areas” (2014 Engel, Pg. 922 (76)), policymakers may feel uneasy about working with providers that legally do not exist but recognise that not doing so may result in lost opportunities. This did not go unrecognised by some practitioners who felt that “[PPM] policy has ignored the potential of alternative systems of medicine and the contribution they can bring to TB control efforts” (2016 Salve, Pg. 631 (42)). The role of external donors was explored in one study where qualitative participants highlighted the role of the leading global stakeholder in supporting programme implementation, knowledge transfer and capacity building (71). However, it was also noted that in the long-term, this proved unsustainable (55, 71).

## Discussion

### Principal findings of the review

In this systematic review, we summarise the impact of PPM models for TB diagnosis and treatment specifically within urban health systems in least developed, low income, and lower-middle-income countries and territories. Overall, we found a positive impact on health outcomes (treatment success, sputum conversion), access to healthcare and coverage (TB diagnostic services, case detection), and value for money (cost-saving interventions, reduced patient costs). We also found that PPM models promoted and improved TB health workers’ skills and service delivery. Any differences in impact based on PPM model type (direct or interface) was less clear though. The interface models were more commonly linked with higher treatment success (close to 90%), reduced TB incidence (by 8.5%), improved awareness and behaviour change among PPs, effective communication, and mutual respect between the public and private sectors. On the other hand, we could not find any pattern of PPM models linked to costs and cost-effectiveness, as just one of ten studies on this outcome was based on a direct model.

Despite the overall positive impact of PPM TB in the urban scenario, our review also pointed out areas of improvement, particularly in the process and system outcomes. GeneXpert testing and TB/HIV co-infection testing were poorer under PPMs compared to the public sector. Qualitative findings reported inconsistencies in the implementation of NTP guidelines, uncoordinated referrals, lack of confidence in the capabilities of partners, inappropriate power dynamics, and inefficiency of direct models when they adopted strict report systems and supervision.

### Interpretation of findings

The WHO recommends PPM models for TB care where there is high utilisation of the private sector, poor quality of care, low case detection, poor treatment outcomes, and increased costs to affected families (90). In the context of LMICs, rapid urbanisation has exacerbated these problems, and our review findings indicate the contribution of PPMs in ameliorating them. Our findings are also supported by previous reviews on the topic (14, 15), although their inclusion criteria covered rural contexts as well. Similar to Lei et al., (15), our included studies were largely from Asia, followed by the African and American Regions. In contrast, whilst they classified the PPM collaborative characteristics as support, contract, and multi-partner groups, we categorised them as direct and interface models and within both model types, listed the roles for public and private sector partners (stewardship/ support/ service provision/ monitoring/ financing), as defined by Tabrizi et al (22). Based on their classification, Lei et al. (15) recommend multiple collaboration mechanisms, including multi-partner groups to ensure positive PPM performance. Although our findings similarly lean towards interface models with multiple partners, this must be interpreted with caution due to the heterogeneity of PPM schemes in terms of coverage, services provided, and outcome measures evaluated.

An earlier review by Malmborg et al. (14) assesses the degree to which the STOP TB Partnership’s (91) global objectives of engaging all care providers are met through existing PPM interventions, and find inclusive evidence on reducing patient costs. Our updated searches identified ten studies reporting cost and/ or cost-effectiveness of PPM models, and findings indicate lower out-of-pocket payments and cost-saving interventions from the societal perspective, possibly due to the influence of lower patient costs despite high implementation costs. However, we found substantial differences in outcomes evaluated, types of costs included, and methodological approaches (e.g., cost-effectiveness thresholds) (Appendix 8), which likely influenced the high degree of variation in implementation costs and whether a model was cost-effective when other study perspectives were adopted. Another 2006 review by Malmborg et al. focuses on the range of PPs included in TB PPMs and conclude that existing models do not adequately cover providers who may be best suited to meet the needs of the poor and vulnerable (92). This is unfortunately still true; few studies mentioned non-allopathic PPs (30, 54, 62, 89), and even fewer reported on informal providers (42, 79), despite existing evidence that all providers can be successfully engaged in TB care with appropriate support and training (93).

Regarding system outcomes, findings from the present review somewhat align with the current evidence. The role of training in improving skills and service delivery within TB PPMs is well documented (15, 94), and challenges related to funding discontinuation and lack of regulatory mechanisms have been noted in non-TB PPMs as well (22, 95). Some recent studies have explored solutions to the system challenges identified such as inconsistent implementation of NTP guidelines, uncoordinated referrals, and lack of trust among partners. A study from Nigeria has recommended making less bulky and more precise NTP guidelines available at all levels of care, as well as training intensification for PPs to improve adherence (96). In another study, a Hub and Spoke model improved the referral system and uptake of GeneXpert testing after a coordinated engagement of private laboratories (97). In India, recognition of PPs as key stakeholders and equal partners, and formalization of partnerships enhanced transparency and trust between partners and a sense of accountability within PPM models (98).

### Strengths and weaknesses

Our review analyses the impact of PPM models for TB diagnosis and treatment, focusing on urban settings in LMICs, to provide decision-makers with systematically synthesised evidence that is specific to these contexts. We excluded upper-middle-income countries, given the differences in trends of urbanisation compared to other LMIC groups (99). Our searches covered a large number of databases and grey literature resources, with no restrictions on outcome, publication date, or language. However, the following are some limitations to consider. First, more than half the studies (65%) were from South-east Asia, with other WHO regions being less represented. So, the extrapolation of our findings may be limited, particularly for South American countries with only one included study. Despite the extensive searches, studies may have been missed. Second, most studies reached a low-quality score (3 or lower), likely reflecting on the accuracy of the information provided. Third, we could not establish a clear pathway of impact according to the PPM model types (direct or interface) due to the heterogeneity of the studies. Finally, we found limited evidence on informal providers, who play an essential role in the TB cascade of care in urban LMIC contexts.

### Implications of findings

Although PPM models have the ability to improve access to TB screening, diagnosis, and treatment outcomes in urban contexts, careful consideration is needed in their design due to considerable levels of mistrust between public and private sectors in most contexts. Factors such as excess control and top-down approaches from government agencies can undermine the strong partnership between public and private providers. Instead, where regular communication built on principles of equal partnership is implemented, PPM models appear to be more successful. There is also some evidence that this may be more achievable when an interface organisation manages the partnership.

Based on ten included studies, PPM interventions appear cost saving, with lower out-of-pocket payments. Nonetheless, more comparative research is needed to understand which PPM models are most cost-effective, particularly to guide the decision-making process and indicate the sustainability of interventions in resource-constrained scenarios (100). Further, Adepoju et al. (101) point out the difficulties in evaluating TB-PPM performances, given the variations in risk profiles, as well as access to providers and services across the public and private sectors in different urban contexts. In addition, we found poor methodological and reporting quality of existing studies. Following from these points, we support the need for clearer and more standard reporting of PPM models and their performances, with particular attention to the involvement of diverse PPs, including informal providers.

## Conclusions

Taken together, our findings support the implementation of PPM models for TB care in urban LMIC contexts. Providing decision-makers with evidence of the best design of PPM models to deliver these positive outcomes is, however, less straightforward. Very few studies included informal providers, which is a major gap in urban LMIC contexts.

## Funding and ethics statements

### Ethical approval

Not applicable.

### Data availability statement

All data analysed during this study are included in this published article and its supplementary information files.

## Acknowledgments

CHORUS funded by UK Aid, from the UK Government, Grant 301132

## Non-author contributors

The authors extend their gratitude to all collaborators for their contributions in data collection and interpretation for this review.

- Dr Bassey Ebenso, UK
- Md Faisal Kabir, Bangladesh
- Deepa Barua, Bangladesh
- Juliana Onuh, Nigeria
- Farzana Sehrin, Bangladesh
- Anna Ray, UK
- Jemima Sumboh, UK
- Nina Amedzro, UK
- Sushil Baral, Nepal

## Contributor and guarantor information

- Conceptualisation: Helen Elsey, Zahidul Quayyum
- Design and development of review protocol: Helen Elsey, Aishwarya Lakshmi Vidyasagaran, Zahidul Quayyum, Baby Naznin, Jannatun Tajree, Mohammad Faisal Kabir, Maisha Ahsan, Deepa Barua, Sampurna Kakchapati, Deepak Joshi, Grishu Shrestha, Florence Tochukwu Sibeudu, Pamela Adaobi Ogbozor, Prince Agwu, Juliana Onuh, Chukwuedozie Ajaero, Chinyere Okeke, Abena Engmann, Bassey Ebenso
- Design of literature search strategy: Su Golder
- Conducting literature searches and data acquisition: Su Golder, Aishwarya Lakshmi Vidyasagaran
- Screening: Aishwarya Lakshmi Vidyasagaran, Helen Elsey, Anna Ray, Zahidul Quayyum, Baby Naznin, Jannatun Tajree, Mohammad Faisal Kabir, Maisha Ahsan, Deepa Barua, Sampurna Kakchapati, Deepak Joshi, Grishu Shrestha, Florence Tochukwu Sibeudu, Pamela Adaobi Ogbozor, Prince Agwu, Juliana Onuh, Bassey Ebenso
- Data extraction and quality appraisal: Aishwarya Lakshmi Vidyasagaran, Helen Elsey, Noemia Siqueira, Jemima Sumboh, Zahidul Quayyum, Baby Naznin, Jannatun Tajree, Swaksar Adhikary, Farzana Sehrin, Mohammad Faisal Kabir, Maisha Ahsan, Deepa Barua, Sampurna Kakchapati, Deepak Joshi, Grishu Shrestha, Florence Tochukwu Sibeudu, Pamela Adaobi Ogbozor
- Data interpretation: Aishwarya Lakshmi Vidyasagaran, Helen Elsey, Noemia Siqueira, Thomas Falconer Hall, Zahidul Quayyum, Baby Naznin, Jannatun Tajree, Sampurna Kakchapati, Deepak Joshi, Grishu Shrestha, Sushil Baral, Ifeyinwa Ngozi Arize
- Manuscript writing: Aishwarya Lakshmi Vidyasagaran, Helen Elsey, Noemia Siqueira, Thomas Falconer Hall, Zahidul Quayyum, Baby Naznin, Jannatun Tajree, Sampurna Kakchapati, Deepak Joshi, Grishu Shrestha, Florence Tochukwu Sibeudu, Pamela Adaobi Ogbozor, Ifeyinwa Ngozi Arize
- Revision of manuscript and editing: All authors critically reviewed and revised the manuscript. All authors had full access to all the data in the study and had final responsibility for the decision to submit for publication.

The corresponding author attests that all listed authors meet authorship criteria and that no others meeting the criteria have been omitted.

## Competing interests declaration

No competing interests.

## Notes

### Competing Interest Statement

The authors have declared no competing interest.

## References

1. World Health Organization. Global tuberculosis report 2021. Geneva: World Health Organization; 2021.

2. Zimmer AJ, Klinton JS, Oga-Omenka C, Heitkamp P, Nyirenda CN, Furin J, et al. Tuberculosis in times of COVID-19. J Epidemiol Community Health. 2021.

3. World Health Organization. The end TB strategy. Geneva: World Health Organization; 2015.

4. Floyd K, Glaziou P, Houben R, Sumner T, White R, Raviglione M. Global tuberculosis targets and milestones set for 2016–2035: definition and rationale. The international journal of tuberculosis and lung disease. 2018;22(7):723–30.

5. Silva S, Arinaminpathy N, Atun R, Goosby E, Reid M. Economic impact of tuberculosis mortality in 120 countries and the cost of not achieving the Sustainable Development Goals tuberculosis targets: a full-income analysis. The Lancet Global Health. 2021;9(10):e1372–e9.

5. Stop TB Partnership. How the partnership works. 2022.

7. Elsey H, Agyepong I, Huque R, Quayyem Z, Baral S, Ebenso B, et al. Rethinking health systems in the context of urbanisation: challenges from four rapidly urbanising low-income and middle-income countries. BMJ global health. 2019;4(3).

8. Adams AM, Islam R, Ahmed T. Who serves the urban poor? A geospatial and descriptive analysis of health services in slum settlements in Dhaka, Bangladesh. Health policy and planning. 2015;30(suppl_1):i32-i45.

9. Mackintosh M, Channon A, Karan A, Selvaraj S, Cavagnero E, Zhao H. What is the private sector? Understanding private provision in the health systems of low-income and middle-income countries. The Lancet. 2016;388(10044):596–605.

10. Lönnroth K, Jaramillo E, Williams BG, Dye C, Raviglione M. Drivers of tuberculosis epidemics: the role of risk factors and social determinants. Social science & medicine. 2009;68(12):2240–6.

11. Arinaminpathy N, Batra D, Maheshwari N, Swaroop K, Sharma L, Sachdeva KS, et al. Tuberculosis treatment in the private healthcare sector in India: an analysis of recent trends and volumes using drug sales data. BMC Infectious Diseases. 2019;19:1–7.

12. Noykhovich E, Mookherji S, Roess A. The risk of tuberculosis among populations living in slum settings: a systematic review and meta-analysis. Journal of Urban Health. 2019;96:262–75.

13. Dewan PK, Lal S, Lonnroth K, Wares F, Uplekar M, Sahu S, et al. Improving tuberculosis control through public-private collaboration in India: literature review. Bmj. 2006;332(7541):574-8.

14. Malmborg R, Mann G, Squire SB. A systematic assessment of the concept and practice of public-private mix for tuberculosis care and control. International journal for equity in health. 2011;10:1–19.

15. Lei X, Liu Q, Escobar E, Philogene J, Zhu H, Wang Y, et al. Public–private mix for tuberculosis care and control: a systematic review. International Journal of Infectious Diseases. 2015;34:20–32.

16. National Academies of Sciences E, Medicine,. Health-Focused Public–Private Partnerships in the Urban Context: Proceedings of a Workshop. 2020.

17. Page MJ, McKenzie JE, Bossuyt PM, Boutron I, Hoffmann TC, Mulrow CD, et al. The PRISMA 2020 statement: an updated guideline for reporting systematic reviews. International journal of surgery. 2021;88:105906.

18. Quayyum Z, Naznin B, Tajree J, Barua D, Ahsan M, Kabir F, et al. Assessment of Public Private Partnership (PPP) models in urban health systems in least developed, low-income and lower middle-income countries and territories: a systematic review. PROSPERO. 2021.

19. Naznin B, Quayyum Z, Tajree J, Golder S, Ebenso B, Barua D, et al. Assessment of Public-Private Partnership (PPP) Models in Health Systems in Least Developed, Low Income and Lower-Middle-Income Countries and Territories: A Protocol for a Systematic Review. Health Care: Current Reviews. 2022.

20. The World Bank. World Bank Country and Lending Groups 2021 [Available from: https://datahelpdesk.worldbank.org/knowledgebase/articles/906519-world-bank-country-and-lending-groups.

21. Hong QN, Fàbregues S, Bartlett G, Boardman F, Cargo M, Dagenais P, et al. The Mixed Methods Appraisal Tool (MMAT) version 2018 for information professionals and researchers. Education for information. 2018;34(4):285–91.

22. Tabrizi JS, Azami-Aghdash S, Gharaee H. Public-private partnership policy in primary health care: a scoping review. Journal of primary care & community health. 2020;11:2150132720943769.

23. World Health Organization. Everybody’s business: strengthening health systems to improve health outcomes: WHO’s framework for action. Geneva: World Health Organization; 2007. Report No.: 9241596074.

24. Campbell M, McKenzie JE, Sowden A, Katikireddi SV, Brennan SE, Ellis S, et al. Synthesis without meta-analysis (SWiM) in systematic reviews: reporting guideline. bmj. 2020;368.

25. QSR International Pty Ltd. NVivo 1.7 2020 [Available from: https://www.qsrinternational.com/nvivo-qualitative-data-analysis-software/home.

26. Glasgow RE, Harden SM, Gaglio B, Rabin B, Smith ML, Porter GC, et al. RE-AIM planning and evaluation framework: adapting to new science and practice with a 20-year review. Frontiers in public health. 2019;7:64.

27. Quy HT, Lan NT, Lonnroth K, Buu TN, Dieu TT, Hai LT. Public-private mix for improved TB control in Ho Chi Minh City, Vietnam: an assessment of its impact on case detection. Int J Tuberc Lung Dis. 2003;7(5):464–71.

28. Quy HT, Lonnroth K, Lan NT, Buu TN. Treatment results among tuberculosis patients treated by private lung specialists involved in a public-private mix project in Vietnam. Int J Tuberc Lung Dis. 2003;7(12):1139–46.

29. Lonnroth K, Uplekar M, Arora VK, Juvekar S, Lan NT, Mwaniki D, et al. Public-private mix for DOTS implementation: what makes it work? Bull World Health Organ. 2004;82(8):580–6.

30. Krishnan A, Kapoor SK. Involvement of private practitioners in tuberculosis control in Ballabgarh, Northern India. International Journal of Tuberculosis and Lung Disease. 2006;10(3):264–9.

31. Lagrada LP, Uehara N, Kawahara K. Analysis of factors of treatment completion in dots health facilities in metro Manila, Philippines: a case-control study. Kekkaku. 2008;83(12):765–72.

32. Ahmed J, Ahmed M, Laghari A, Lohana W, Ali S, Fatmi Z. Public private mix model in enhancing tuberculosis case detection in District Thatta, Sindh, Pakistan. JPMA J Pak Med Assoc. 2009;59(2):82–6.

33. Pantoja A, Floyd K, Unnikrishnan KP, Jitendra R, Padma MR, Lal SS, et al. Economic evaluation of public-private mix for tuberculosis care and control, India. Part I. Socio-economic profile and costs among tuberculosis patients. Int J Tuberc Lung Dis. 2009;13(6):698–704.

34. Pantoja A, Lonnroth K, Lal SS, Chauhan LS, Uplekar M, Padma MR, et al. Economic evaluation of public-private mix for tuberculosis care and control, India. Part II. Cost and cost-effectiveness. Int J Tuberc Lung Dis. 2009;13(6):705–12.

35. Quelapio MI, Mira NR, Orillaza-Chi RB, Belen V, Munez N, Belchez R, et al. Responding to the multidrug-resistant tuberculosis crisis: mainstreaming programmatic management to the Philippine National Tuberculosis Programme. Int J Tuberc Lung Dis. 2010;14(6):751–7.

36. Lal SS, Sahu S, Wares F, Lonnroth K, Chauhan LS, Uplekar M. Intensified scale-up of public-private mix: a systems approach to tuberculosis care and control in India. Int J Tuberc Lung Dis. 2011;15(1):97–104.

37. Pradhan A, Datye V, Kielmann K, Khilare K, Datye A, Inamdar V, et al. Sustaining PPM-DOTS: the case of Pimpri Chinchwad, Maharashtra, India. Indian J. 2011;58(1):18–28.

38. Daboer JC, Lar LA, Afolaranmi TO, Bupwatda PW, Dani N. Public-private mix in tuberculosis control: an assessment of level of implementation in Jos, Plateau State. Niger Postgrad Med J. 2013;20(4):282–5.

39. Daniel OJ, Adedeji Adejumo O, Abdur-Razzaq HA, Ngozi Adejumo E, Salako AA. Public-private mix for TB and TB-HIV care in Lagos, Nigeria. Int J Tuberc Lung Dis. 2013;17(9):1195–8.

40. Kielmann K, Datye V, Pradhan A, Rangan S. Balancing authority, deference and trust across the public-private divide in health care: tuberculosis health visitors in western Maharashtra, India. Glob Public Health. 2014;9(8):975–92.

41. Subramaniyam S, Chadha VK, Manuvel C, Praseeja P, Sharada MA, Nagendra N, et al. Treatment outcome of tuberculosis patients in a clinic of Bangalore. Indian Journal of Tuberculosis. 2014;61(3):189–94.

42. Salve S, Sheikh K, Porter JD. Private Practitioners’ Perspectives on Their Involvement With the Tuberculosis Control Programme in a Southern Indian State. Int. 2016;5(11):631–42.

43. Khan BJ, Kumar AMV, Stewart A, Khan NM, Selvaraj K, Fatima R, et al. Alarming rates of attrition among tuberculosis patients in public-private facilities in Lahore, Pakistan. Public health action. 2017;7(2):127–33.

44. Qader G, Hamim A, Sayedi M, Rashidi M, Manzoor L, Seddiq MK, et al. Addressing tuberculosis control in fragile states: Urban DOTS experience in Kabul, Afghanistan, 2009-2015. PLoS ONE. 2017;12(5).

45. Reviono R, Setianingsih W, Damayanti KE, Ekasari R. The dynamic of tuberculosis case finding in the era of the public-private mix strategy for tuberculosis control in Central Java, Indonesia. Glob Health Action. 2017;10(1):1353777-.

46. Awan WM, Zaidi SMA, Habib SS, Khowaja S, Malik A, Khan U, et al. Impact of scaling up Xpert (R) MTB/RIF testing for the detection of rifampicin-resistant TB cases in Karachi, Pakistan. Int J Tuberc Lung Dis. 2018;22(8):899-+.

47. Yellappa V, Battaglioli T, Gurum SK, Narayanan D, Van der Stuyft P. Involving private practitioners in the Indian tuberculosis programme: a randomised trial. Tropical Medicine & International Health. 2018;23(5):570–9.

48. Hadisoemarto. Increasing Notifications of Tuberculosis From Private Practitioners. https://clinicaltrialsgov/show/NCT04187313. 2019.

49. Hadisoemarto PF, Lestari BW, Sharples K, Afifah N, Chaidir L, Huang C-C, et al. A public health intervention package for increasing tuberculosis notifications from private practitioners in Bandung, Indonesia (INSTEP2): A cluster-randomised controlled trial protocol. F1000Research. 2022;10:327-.

50. Hemavarneshwari S, Shaikh RB, Naik PR, Nagaraja SB. Strategy to sensitize private practitioners on RNTCP through medico-social workers in urban field practice area of a Medical College in Bengaluru, Karnataka. Indian J. 2019;66(2):253–8.

51. Paul KK, Alkabab YMA, Rahman MM, Ahmed S, Amin MJ, Hossain MD, et al. A public-private model to scale up diabetes mellitus screening among people accessing tuberculosis diagnostics in Dhaka, Bangladesh. International Journal of Infectious Diseases. 2020;92:56–61.

52. Thu TD, Kumar AMV, Ramaswamy G, Htun T, Van HL, Quang LVN, et al. An Innovative Public-Private Mix Model for Improving Tuberculosis Care in Vietnam: How Well are We Doing? Trop. 2020;5(1):14-.

53. Oladimeji O, Adepoju V, Anyiam FE, San JE, Odugbemi BA, Hyera FLM, et al. Treatment outcomes of drug susceptible Tuberculosis in private health facilities in Lagos, South-West Nigeria. PLoS ONE. 2021;16(1):e0244581.

54. Murthy KJ, Frieden TR, Yazdani A, Hreshikesh P. Public-private partnership in tuberculosis control: experience in Hyderabad, India. Int J Tuberc Lung Dis. 2001;5(4):354–9.

55. Hurtig AK, Pande SB, Baral SC, Newell J, Porter JDH, Bam DS. Linking private and public sectors in tuberculosis treatment in Kathmandu Valley, Nepal. Health policy and planning. 2002;17(1):78–89.

56. Rangan S, Ambe G, Borremans N, Zallocco D, Porter J. The Mumbai experience in building field level partnerships for DOTS implementation. Tuberculosis. 2003;83(1-3):165–72.

57. Tupasi TE, Quelapio MI, Orillaza RB, Alcantara C, Mira NR, Abeleda MR, et al. DOTS-Plus for multidrug-resistant tuberculosis in the Philippines: global assistance urgently needed. Tuberculosis (Edinb). 2003;83(1-3):52–8.

58. Tupasi TE, Gupta R, Quelapio MID, Orillaza RB, Mira NR, Mangubat NV, et al. Feasibility and Cost-Effectiveness of Treating Multidrug-Resistant Tuberculosis: A Cohort Study in the Philippines. PLoS Medicine. 2006;3(9):e352-e.

59. Arora VK, Lonnroth K, Sarin R. Improved case detection of tuberculosis through a public-private partnership. Indian J Chest Dis Allied Sci. 2004;46(2):133–6.

60. Newell JN, Pande SB, Baral SC, Bam DS, Malla P. Control of tuberculosis in an urban setting in Nepal: public-private partnership. Bull World Health Organ. 2004;82(2):92–8.

61. Newell JN, Pande SB, Baral SC, Bam DS, Malla P. Leadership, management and technical lessons learnt from a successful public-private partnership for TB control in Nepal. Int J Tuberc Lung Dis. 2005;9(9):1013–7.

62. Ambe G, Lonnroth K, Dholakia Y, Copreaux J, Zignol M, Borremans N, et al. Every provider counts: effect of a comprehensive public-private mix approach for TB control in a large metropolitan area in India. Int J Tuberc Lung Dis. 2005;9(5):562–8.

63. Lambert ML, Delgado R, Michaux G, Vols A, Speybroeck N, Van der Stuyft P. Collaboration between private pharmacies and national tuberculosis programme: an intervention in Bolivia. Trop Med Int Health. 2005;10(3):246–50.

64. Floyd K, Arora VK, Murthy KJ, Lonnroth K, Singla N, Akbar Y, et al. Cost and cost-effectiveness of PPM-DOTS for tuberculosis control: evidence from India. Bull World Health Organ. 2006;84(6):437–45.

65. Maung M, Kluge H, Aye T, Maung W, Noe P, Zaw M, et al. Private GPs contribute to TB control in Myanmar: evaluation of a PPM initiative in Mandalay Division. Int J Tuberc Lung Dis. 2006;10(9):982–7.

66. Irawati SR, Basri C, Arias MS, Prihatini S, Rintiswati N, Voskens J, et al. Hospital DOTS linkage in Indonesia: a model for DOTS expansion into government and private hospitals. Int J Tuberc Lung Dis. 2007;11(1):33–9.

67. Karki DK, Mirzoev TN, Green AT, Newell JN, Baral SC. Costs of a successful public-private partnership for TB control in an urban setting in Nepal. BMC Public Health. 2007;7:84-.

68. Chakaya J, Uplekar M, Mansoer J, Kutwa A, Karanja G, Ombeka V, et al. Public-private mix for control of tuberculosis and TB-HIV in Nairobi, Kenya: outcomes, opportunities and obstacles. Int J Tuberc Lung Dis. 2008;12(11):1274–8.

69. Krishnan N, Ananthakrishnan R, Augustine S, Vijayalakshmi NK, Gopi PG, Kumaraswami V, et al. Impact of advocacy on the tuberculosis management practices of private practitioners in Chennai City, India. Int J Tuberc Lung Dis. 2009;13(1):112–8.

70. Bell CA, Eang MT, Dareth M, Rothmony E, Duncan GJ, Saini B. Provider perceptions of pharmacy-initiated tuberculosis referral services in Cambodia, 2005-2010. Int J Tuberc Lung Dis. 2012;16(8):1086-91.

71. Bell CA, Duncan GJ, Eang R, Saini B. Stakeholder perceptions of a pharmacy-initiated tuberculosis referral program in Cambodia, 2005-2012. Asia Pac J Public Health. 2015;27(2):NP2570–7.

72. Khan AJ, Khowaja S, Khan FS, Qazi F, Lotia I, Habib A, et al. Engaging the private sector to increase tuberculosis case detection: an impact evaluation study. Lancet Infect Dis. 2012;12(8):608–16.

73. Naqvi SA, Naseer M, Kazi A, Pethani A, Naeem I, Zainab S, et al. Implementing a public-private mix model for tuberculosis treatment in urban Pakistan: lessons and experiences. Int J Tuberc Lung Dis. 2012;16(6):817–21.

74. Zafar Ullah AN, Huque R, Husain A, Akter S, Islam A, Newell JN. Effectiveness of involving the private medical sector in the National TB Control Programme in Bangladesh: evidence from mixed methods. BMJ Open. 2012;2(6).

75. Zafar Ullah AN, Huque R, Husain A, Akter S, Akter H, Newell JN. Tuberculosis in the workplace: developing partnerships with the garment industries in Bangladesh. Int J Tuberc Lung Dis. 2012;16(12):1637–42.

76. Engel N, van Lente H. Organisational innovation and control practices: the case of public-private mix in tuberculosis control in India. Sociol Health Illn. 2014;36(6):917–31.

77. Pethani A, Zafar M, Khan AA, Rabbani U, Ahmed S, Fatmi Z. Engaging general practitioners in public-private mix tuberculosis DOTS program in an urban area in Pakistan: need for context-specific approach. Asia Pac J Public Health. 2015;27(2):NP984–92.

78. Ramaiah AA, Gawde NC. Economic Evaluation of a Public-Private Mix TB Project in Tamil Nadu, India. J Health Manag. 2015;17(3):370–80.

79. Bronner Murrison L, Ananthakrishnan R, Swaminathan A, Auguesteen S, Krishnan N, Pai M, et al. How do patients access the private sector in Chennai, India? An evaluation of delays in tuberculosis diagnosis. Int J Tuberc Lung Dis. 2016;20(4):544–51.

80. Lestari BW, Arisanti N, Siregar AYM, Sihaloho ED, Budiman G, Hill PC, et al. Feasibility study of strengthening the public-private partnership for tuberculosis case detection in Bandung City, Indonesia. BMC Res Notes. 2017;10(1):404-.

81. Chadha VK, Bhalla BB, Ramesh SB, Gupta J, Nagendra N, Padmesh R, et al. Tuberculosis diagnostic and treatment practices in private sector: Implementation study in an Indian city. Indian J. 2018;65(4):315–21.

82. McDowell A, Raizada N, Khaparde SD, Rao R, Sarin S, Kalra A, et al. "Before Xpert I only had my expertise": A qualitative study on the utilization and effects of Xpert technology among pediatricians in 4 Indian cities. PLoS ONE. 2018;13(3).

83. Ananthakrishnan R, Richardson MD, van den Hof S, Rangaswamy R, Thiagesan R, Auguesteen S, et al. Successfully Engaging Private Providers to Improve Diagnosis, Notification, and Treatment of TB and Drug-Resistant TB: The EQUIP Public-Private Model in Chennai, India. Glob. 2019;7(1):41–53.

84. Arinaminpathy N, Mandal S, Bhatia V, McLeod R, Sharma M, Swaminathan S, et al. Strategies for ending tuberculosis in the South-East Asian Region: A modelling approach. Indian Journal of Medical Research. 2019;149(4):517-.

85. Arinaminpathy N, Nandi A, Vijayan S, Jha N, Nair SA, Kumta S, et al. Engaging with the private healthcare sector for the control of tuberculosis in India: cost and cost-effectiveness. Bmj Global Health. 2021;6(10).

86. Daftary A, Satyanarayana S, Jha N, Singh M, Mondal S, Vadnais C, et al. Can community pharmacists improve tuberculosis case finding? A mixed methods intervention study in India. BMJ glob. 2019;4(3):e001417.

87. Ramasamy R, Mohanta GP, Hiremath SRR, Dang R, Chandramouli R, Gharat MS. Assessing the change of community pharmacist’s knowledge on tuberculosis and attitude to practice as a tuberculosis dots provider after an educational intervention. International Journal of Research in Pharmaceutical Sciences. 2020;11(4):7593–9.

88. Ramasamy R, Mohanta GP, Hiremath SRR, Ramnarayanan C, Dang R, Gharat MS. Dynamic method for liaison of community pharmacists with national programme for tuberculosis control: Efforts to harness untapped opportunities. Indian Journal of Pharmaceutical Education and Research. 2020;54(3):809–18.

89. Shibu V, Daksha S, Rishabh C, Sunil K, Devesh G, Lal S, et al. Tapping private health sector for public health program? Findings of a novel intervention to tackle TB in Mumbai, India. Indian J. 2020;67(2):189–201.

90. World Health Organization. Tuberculosis: Public-private mix (PPM) for TB care and control Q&A. 2015.

91. World Health Organization TSTP. The Stop TB Strategy: building on and enhancing DOTS to meet the TB-related Millennium Development Goals. Geneva: World Health Organization; 2006.

92. Malmborg R, Mann G, Thomson R, Squire SB. Can public-private collaboration promote tuberculosis case detection among the poor and vulnerable? Bulletin of the World Health Organization. 2006;84:752–8.

93. Thapa P, Jayasuriya R, Hall JJ, Beek K, Mukherjee P, Gudi N, et al. Role of informal healthcare providers in tuberculosis care in low-and middle-income countries: A systematic scoping review. Plos one. 2021;16(9):e0256795.

94. Amo-Adjei J. Conforming to partnership values: a qualitative case study of public–private mix for TB control in Ghana. Global Health Action. 2016;9(1):28000.

95. Basabih M, Prasojo E, Rahayu AYS. Hospital services under public-private partnerships, outcomes and, challenges: A literature review. Journal of Public Health Research. 2022;11(3):22799036221115781.

96. Adepoju VA, Adejumo OA, Adepoju OE, Adeniyi MO, Etuk V, Nzekwe I, et al. Do private health providers adhere to National Tuberculosis Guideline while assigning treatment outcome? Findings from a lower middle-income country. Frontiers in Public Health. 2022;10:924132.

97. Ali T, Singh U, Ohikhuai C, Panwal T, Adetiba T, Agbaje A, et al. Partnering with the private laboratories to strengthen TB diagnostics in Nigeria. Journal of Clinical Tuberculosis and Other Mycobacterial Diseases. 2023:100369.

98. Anand T, Babu R, Jacob AG, Sagili K, Chadha SS. Enhancing the role of private practitioners in tuberculosis prevention and care activities in India. Lung India: Official Organ of Indian Chest Society. 2017;34(6):538.

99. Ritchie H, Roser M. Urbanization. Our World in Data. 2018.

100. Slevin K, Forbes A, Wells W. Public Private Mix (PPM) Models for the Sustainability of Successful TB Control Initiatives. A working meeting co-convened by USAID and the World Bank, in collaboration with the Stop TB Partnership’s PPM subgroup, and organized with PATH Washington, DC: USAID, The World Bank; 2014.

101. Adepoju VA, Oladimeji O, Horsburgh CR, editors. Rethinking public private mix (PPM) performance in the tuberculosis program: how is care seeking impacting this model in high TB burden countries? Healthcare; 2022: MDPI.

