## Appendices 1-8 for "Public–Private Mix (PPM) for Tuberculosis (TB) in Urban Health Systems in Least Developed, Low Income and Lower-Middle-Income Countries and Territories – A Systematic Review"

### Supplementary material

#### Appendix 1: PRISMA Checklist[1]

| Section and Topic | Item # | Checklist item | Location where item is reported |
|---|---|---|---|
| **TITLE** | | | |
| Title | 1 | Identify the report as a systematic review. | Pg. 1 |
| **ABSTRACT** | | | |
| Abstract | 2 | See the PRISMA 2020 for Abstracts checklist (below) | Next table |
| **INTRODUCTION** | | | |
| Rationale | 3 | Describe the rationale for the review in the context of existing knowledge. | Pgs. 4 and 5 |
| Objectives | 4 | Provide an explicit statement of the objective(s) or question(s) the review addresses. | Abstract and Pg. 5 |
| **METHODS** | | | |
| Eligibility criteria | 5 | Specify the inclusion and exclusion criteria for the review and how studies were grouped for the syntheses. | Pg. 6 (also appendix 3) |
| Information sources | 6 | Specify all databases, registers, websites, organisations, reference lists and other sources searched or consulted to identify studies. Specify the date when each source was last searched or consulted. | Pg. 5 (also appendix 2) |
| Search strategy | 7 | Present the full search strategies for all databases, registers, and websites, including any filters and limits used. | Appendix 2 |
| Selection process | 8 | Specify the methods used to decide whether a study met the inclusion criteria of the review, including how many reviewers screened each record and each report retrieved, whether they worked independently, and if applicable, details of automation tools used in the process. | Pg. 6 |
| Data collection process | 9 | Specify the methods used to collect data from reports, including how many reviewers collected data from each report, whether they worked independently, any processes for obtaining or confirming data from study investigators, and if applicable, details of automation tools used in the process. | Pg. 6 |
| Data items | 10a | List and define all outcomes for which data were sought. Specify whether all results that were compatible with each outcome domain in each study were sought (e.g., for all measures, time points, analyses), and if not, the methods used to decide which results to collect. | Pg. 6 |
| | 10b | List and define all other variables for which data were sought (e.g., participant and intervention characteristics, funding sources). Describe any assumptions made about any missing or unclear information. | Pg. 6 |
| Study risk of bias assessment | 11 | Specify the methods used to assess risk of bias in the included studies, including details of the tool(s) used, how many reviewers assessed each study and whether they worked independently, and if applicable, details of automation tools used in the process. | Pg. 6 |
| Effect measures | 12 | Specify for each outcome the effect measure(s) (e.g., risk ratio, mean difference) used in the synthesis or presentation of results. | Pg. 6 |

[1] From: Page MJ, McKenzie JE, Bossuyt PM, Boutron I, Hoffmann TC, Mulrow CD, et al. The PRISMA 2020 statement: an updated guideline for reporting systematic reviews. BMJ 2021;372:n71. doi: 10.1136/bmj.n71

| Section and Topic | Item # | Checklist item | Location where item is reported |
|---|---|---|---|
| Synthesis methods | 13a | Describe the processes used to decide which studies were eligible for each synthesis (e.g., tabulating the study intervention characteristics and comparing against the planned groups for each synthesis (item #5)). | Pgs. 6 and 7 |
| | 13b | Describe any methods required to prepare the data for presentation or synthesis, such as handling of missing summary statistics, or data conversions. | Pgs. 6 and 7 |
| | 13c | Describe any methods used to tabulate or visually display results of individual studies and syntheses. | Pgs. 6 and 7 |
| | 13d | Describe any methods used to synthesize results and provide a rationale for the choice(s). If meta-analysis was performed, describe the model(s), method(s) to identify the presence and extent of statistical heterogeneity, and software package(s) used. | Pgs. 6 and 7 |
| | 13e | Describe any methods used to explore possible causes of heterogeneity among study results (e.g. subgroup analysis, meta-regression). | NA |
| | 13f | Describe any sensitivity analyses conducted to assess robustness of the results. | NA |
| Reporting bias assessment | 14 | Describe any methods used to assess risk of bias due to missing results in a synthesis (arising from reporting biases). | NA |
| Certainty assessment | 15 | Describe any methods used to assess certainty (or confidence) in the body of evidence for an outcome. | NA |
| **RESULTS** | | | |
| Study selection | 16a | Describe the results of the search and selection process, from the number of records identified in the search to the number of studies included in the review, ideally using a flow diagram. | Pg. 6 |
| | 16b | Cite studies that might appear to meet the inclusion criteria, but which were excluded, and explain why they were excluded. | Pg. 6 (also appendix 5) |
| Study characteristics | 17 | Cite each included study and present its characteristics. | Pg. 8 and Table 1 (also appendix 5) |
| Risk of bias in studies | 18 | Present assessments of risk of bias for each included study. | Pg. 8 and Table 1 (also appendix 5) |
| Results of individual studies | 19 | For all outcomes, present, for each study: (a) summary statistics for each group (where appropriate) and (b) an effect estimate and its precision (e.g. confidence/credible interval), ideally using structured tables or plots. | Table 1; also Pgs. 17-20 |
| Results of syntheses | 20a | For each synthesis, briefly summarise the characteristics and risk of bias among contributing studies. | Pgs. 8 and 22 |
| | 20b | Present results of all statistical syntheses conducted. If meta-analysis was done, present for each the summary estimate and its precision (e.g. confidence/credible interval) and measures of statistical heterogeneity. If comparing groups, describe the direction of the effect. | Pgs. 22 and 23 |
| | 20c | Present results of all investigations of possible causes of heterogeneity among study results. | Pg. 22 (also appendix 9) |
| | 20d | Present results of all sensitivity analyses conducted to assess the robustness of the synthesized results. | NA |
| Reporting biases | 21 | Present assessments of risk of bias due to missing results (arising from reporting biases) for each synthesis assessed. | Pgs. 22 and 23 (also appendix 9) |
| Certainty of evidence | 22 | Present assessments of certainty (or confidence) in the body of evidence for each outcome assessed. | NA |
| **DISCUSSION** | | | |
| Discussion | 23a | Provide a general interpretation of the results in the context of other evidence. | Pgs. 23 and 24 |
| | 23b | Discuss any limitations of the evidence included in the review. | Pgs. 24 and 25 |
| | 23c | Discuss any limitations of the review processes used. | Pg. 25 |
| | 23d | Discuss implications of the results for practice, policy, and future research. | Pg. 25 |
| **OTHER INFORMATION** | | | |
| Registration and protocol | 24a | Provide registration information for the review, including register name and registration number, or state that the review was not registered. | Pg. 4 |
| | 24b | Indicate where the review protocol can be accessed, or state that a protocol was not prepared. | PROSPERO |
| | 24c | Describe and explain any amendments to information provided at registration or in the protocol. | Pg. 24 |

| Section and Topic | Item # | Checklist item | Location where item is reported |
|---|---|---|---|
| Support | 25 | Describe sources of financial or non-financial support for the review, and the role of the funders or sponsors in the review. | Abstract, Pgs. 6 and 26 |
| Competing interests | 26 | Declare any competing interests of review authors. | Pg. 26 |
| Availability of data, code and other materials | 27 | Report which of the following are publicly available and where they can be found: template data collection forms; data extracted from included studies; data used for all analyses; analytic code; any other materials used in the review. | Extracted data – Tables and appendices |

| Section and Topic | Item # | Checklist item | Reported (yes/no) |
|---|---|---|---|
| **TITLE** | | | |
| Title | 1 | Identify the report as a systematic review. | Y |
| **BACKGROUND** | | | |
| Objectives | 2 | Provide an explicit statement of the main objective(s) or question(s) the review addresses. | Y |
| **METHODS** | | | |
| Eligibility criteria | 3 | Specify the inclusion and exclusion criteria for the review. | Y |
| Information sources | 4 | Specify the information sources (e.g. databases, registers) used to identify studies and the date when each was last searched. | N |
| Risk of bias | 5 | Specify the methods used to assess risk of bias in the included studies. | Y |
| Synthesis of results | 6 | Specify the methods used to present and synthesise results. | Y |
| **RESULTS** | | | |
| Included studies | 7 | Give the total number of included studies and participants and summarise relevant characteristics of studies. | Y |
| Synthesis of results | 8 | Present results for main outcomes, preferably indicating the number of included studies and participants for each. If meta-analysis was done, report the summary estimate and confidence/credible interval. If comparing groups, indicate the direction of the effect (i.e. which group is favoured). | Y |
| **DISCUSSION** | | | |
| Limitations of evidence | 9 | Provide a brief summary of the limitations of the evidence included in the review (e.g. study risk of bias, inconsistency and imprecision). | Y |
| Interpretation | 10 | Provide a general interpretation of the results and important implications. | Y |
| **OTHER INFORMATION** | | | |
| Funding | 11 | Specify the primary source of funding for the review. | Y |
| Registration | 12 | Provide the register name and registration number. | Y |

### Appendix 2: Databases & repositories searched and search strategies

| Database or repository name | Database/ repository platform/ URL |
|---|---|
| MEDLINE(R) ALL<br><1946 to August 12, 2022> | Ovid |
| Embase Classic+Embase<br><1974 to 2022 August 12> | Ovid |
| HMIC Health Management Information Consortium <1979 to August 2022> | Ovid |
| Science Citation Index-Expanded (Web of Science) 1900-present<br>Social Sciences Citation Index (Web of Science) 1900-present<br>Emerging Sources Citation Index (Web of Science) 2015-present | Web of Science |
| CENTRAL (Includes EPOC Search Register) | The Cochrane Library |

| | |
|---|---|
| Database of disability and inclusion information resources | https://asksource.info/resources/search? |
| WHOLIS | https://kohahq.searo.who.int/ |
| 3ie International Initiative for Impact Evaluation | https://www.3ieimpact.org/ |

### Ovid MEDLINE(R) ALL 1946 to August 12, 2022

Search date: 15/08/2022; Records: 1,415

- 1 Public-Private Sector Partnerships/ 2436
- 2 (Public adj3 private adj3 partnership$).mp. 4153
- 3 (Public adj3 private adj3 mix$).mp. 576
- 4 (Private adj3 public adj3 collaborat$).mp. 451
- 5 (Private adj3 public adj3 cooperati$).mp. 108
- 6 (Private adj3 public adj3 co-operati$).mp. 10
- 7 (private adj3 public adj3 alliance$).mp. 32
- 8 (private adj3 public adj3 association$).mp. 54
- 9 (private adj3 public adj3 coalition$).mp. 25
- 10 (private adj3 public adj3 union$).mp. 12
- 11 (private adj3 public adj3 consortium$).mp. 49
- 12 (private adj3 public adj3 initiative$).mp. 226
- 13 (private adj3 public adj3 joint$).mp. 46
- 14 (private adj3 public adj3 ownership$).mp. 94
- 15 privatepublic.mp. 5
- 16 publicprivate.mp. 7
- 17 (afghanistan or angola or armenia or armenian or bangladesh or benin or bhutan or bolivia or burkina faso or burkina fasso or upper volta or burundi or urundi or cabo verde or cape verde or cambodia or kampuchea or khmer republic or cameroon or cameron or cameroun or central african republic or ubangi shari or chad or comoros or comoro islands or iles comores or mayotte or democratic republic of the congo or democratic republic congo or congo or zaire or "cote d'ivoire" or "cote d' ivoire" or cote divoire or cote d ivoire or ivory coast or djibouti or french somaliland or egypt or united arab republic or el salvador or eritrea or eswatini or swaziland or ethiopia or gambia or "georgia (republic)" or georgian or ghana or guam or guatemala or guinea or guinea bissau or haiti or hispaniola or honduras or india or indonesia or timor or jordan or kenya or Kiribati or "democratic people's republic of korea" or republic of korea or north korea or korea or kosovo or kyrgyzstan or kirghizia or kirgizstan or kyrgyz republic or kirghiz or laos or lao pdr or "lao people's democratic republic" or lesotho or basutoland or liberia or madagascar or malagasy republic or malawi or nyasaland or mali or micronesia or federated states of micronesia or kiribati or tuvalu or mauritania or moldova or moldovian or mongolia or morocco or ifni or mozambique or portuguese east africa or myanmar or burma or nepal or nicaragua or niger or nigeria or pakistan or papua new guinea or new guinea or philippines or philipines or phillipines or phillippines or rwanda or ruanda or "sao tome and principe" or senegal or sierra leone or melanesia or solomon island or solomon islands or somalia or south sudan or sri lanka or ceylon or sudan or syria or syrian arab republic or tajikistan or tadjikistan or tadzhikistan or tadzhik or tanzania or tanganyika or timor leste or east timor or togo or togolese republic or Tokelau or tunisia or uganda or ukraine or uzbekistan or uzbek or vanuatu or new hebrides

or vietnam or viet nam or middle east or west bank or gaza or palestine or yemen or zambia or zimbabwe or northern rhodesia or global south or africa south of the sahara or sub-saharan africa or subsaharan africa or africa, central or central africa or africa, northern or north africa or northern africa or magreb or maghrib or sahara or africa, southern or southern africa or africa, eastern or east africa or eastern africa or africa, western or west africa or western africa or west indies or indian ocean islands or caribbean or central america or latin america or "south and central america" or south america or asia, central or central asia or asia, northern or north asia or northern asia or asia, southeastern or southeastern asia or south eastern asia or southeast asia or south east asia or asia, western or western asia or europe, eastern or east europe or eastern europe or developing country or developing countries or developing nation? or developing population? or developing world or less developed countr* or least developed nation? or least developed population? or least developed world or least developed countr* or less developed nation? or less developed population? or less developed world or lesser developed countr* or lesser developed nation? or lesser developed population? or lesser developed world or under developed countr* or under developed nation? or under developed population? or under developed world or underdeveloped countr* or underdeveloped nation? or underdeveloped population? or underdeveloped world or lower middle income countr* or lower middle income nation? or lower middle income population? or low income countr* or low income nation? or low income population? or lower income countr* or lower income nation? or lower income population? or underserved countr* or underserved nation? or underserved population? or underserved world or under served countr* or under served nation? or under served population? or under served world or deprived countr* or deprived nation? or deprived population? or deprived world or poor countr* or poor nation? or poor population? or poor world or poorer countr* or poorer nation? or poorer population? or poorer world or developing econom* or less developed econom* or lesser developed econom* or under developed econom* or underdeveloped econom* or lower middle income econom* or low income econom* or lower income econom* or low gdp or low gnp or low gross domestic or low gross national or lower gdp or lower gnp or lower gross domestic or lower gross national or lmic or lmics or third world or lami countr* or transitional countr* or emerging economies or emerging nation?).ti,ab,sh,kf.
1085533

- 18 or/1-16 5464
- 19 17 and 18 1460
- 20 19 not (editorial or letter or comment).pt. 1415

### Ovid Embase <1974 to 2022 August 12>

Date Searched: 15/08/22; Records: 2,034

- 1 Public-Private Sector Partnerships/ 6670
- 2 (Public adj3 private adj3 partnership$).mp. 8181
- 3 (Public adj3 private adj3 mix$).mp. 704
- 4 (Private adj3 public adj3 collaborat$).mp. 543
- 5 (Private adj3 public adj3 cooperati$).mp. 152
- 6 (Private adj3 public adj3 co-operati$).mp. 11
- 7 (private adj3 public adj3 alliance$).mp. 39
- 8 (private adj3 public adj3 association$).mp. 60
- 9 (private adj3 public adj3 coalition$).mp. 37
- 10 (private adj3 public adj3 union$).mp. 16

- 11 (private adj3 public adj3 consortium$).mp. 85
- 12 (private adj3 public adj3 initiative$).mp. 280
- 13 (private adj3 public adj3 joint$).mp. 54
- 14 (private adj3 public adj3 ownership$).mp. 103
- 15 privatepublic.mp. 11
- 16 publicprivate.mp. 53
- 17 (afghanistan or angola or armenia or armenian or bangladesh or benin or bhutan or bolivia or burkina faso or burkina fasso or upper volta or burundi or urundi or cabo verde or cape verde or cambodia or kampuchea or khmer republic or cameroon or cameron or cameroun or central african republic or ubangi shari or chad or comoros or comoro islands or iles comores or mayotte or democratic republic of the congo or democratic republic congo or congo or zaire or "cote d'ivoire" or "cote d' ivoire" or cote divoire or cote d ivoire or ivory coast or djibouti or french somaliland or egypt or united arab republic or el salvador or eritrea or eswatini or swaziland or ethiopia or gambia or "georgia (republic)" or georgian or ghana or guam or guatemala or guinea or guinea bissau or haiti or hispaniola or honduras or india or indonesia or timor or jordan or kenya or Kiribati or "democratic people's republic of korea" or republic of korea or north korea or korea or kosovo or kyrgyzstan or kirghizia or kirgizstan or kyrgyz republic or kirghiz or laos or lao pdr or "lao people's democratic republic" or lesotho or basutoland or liberia or madagascar or malagasy republic or malawi or nyasaland or mali or micronesia or federated states of micronesia or kiribati or tuvalu or mauritania or moldova or moldovian or mongolia or morocco or ifni or mozambique or portuguese east africa or myanmar or burma or nepal or nicaragua or niger or nigeria or pakistan or papua new guinea or new guinea or philippines or philipines or phillipines or phillippines or rwanda or ruanda or "sao tome and principe" or senegal or sierra leone or melanesia or solomon island or solomon islands or somalia or south sudan or sri lanka or ceylon or sudan or syria or syrian arab republic or tajikistan or tadjikistan or tadzhikistan or tadzhik or tanzania or tanganyika or timor leste or east timor or togo or togolese republic or Tokelau or tunisia or uganda or ukraine or uzbekistan or uzbek or vanuatu or new hebrides or vietnam or viet nam or middle east or west bank or gaza or palestine or yemen or zambia or zimbabwe or northern rhodesia or global south or africa south of the sahara or sub-saharan africa or subsaharan africa or africa, central or central africa or africa, northern or north africa or northern africa or magreb or maghrib or sahara or africa, southern or southern africa or africa, eastern or east africa or eastern africa or africa, western or west africa or western africa or west indies or indian ocean islands or caribbean or central america or latin america or "south and central america" or south america or asia, central or central asia or asia, northern or north asia or northern asia or asia, southeastern or southeastern asia or south eastern asia or southeast asia or south east asia or asia, western or western asia or europe, eastern or east europe or eastern europe or developing country or developing countries or developing nation? or developing population? or developing world or less developed countr* or least developed nation? or least developed population? or least developed world or least developed countr* or less developed nation? or less developed population? or less developed world or lesser developed countr* or lesser developed nation? or lesser developed population? or lesser developed world or under developed countr* or under developed nation? or under developed population? or under developed world or underdeveloped countr* or underdeveloped nation? or underdeveloped population? or underdeveloped world or lower middle income countr* or lower middle income nation? or lower middle income population? or low income countr* or low income nation? or low income population? or lower income countr* or lower income nation? or

lower income population? or underserved countr* or underserved nation? or underserved population? or underserved world or under served countr* or under served nation? or under served population? or under served world or deprived countr* or deprived nation? or deprived population? or deprived world or poor countr* or poor nation? or poor population? or poor world or poorer countr* or poorer nation? or poorer population? or poorer world or developing econom* or less developed econom* or lesser developed econom* or under developed econom* or underdeveloped econom* or lower middle income econom* or low income econom* or lower income econom* or low gdp or low gnp or low gross domestic or low gross national or lower gdp or lower gnp or lower gross domestic or lower gross national or lmic or lmics or third world or lami countr* or transitional countr* or emerging economies or emerging nation?).ti,ab,sh,kf.
1236825

- 18 or/1-16 9711
- 19 17 and 18 2347
- 20 19 not (editorial or letter or comment).pt. 2034

### Ovid HMIC Health Management Information Consortium <1979 to August 2022>

Date Searched: 15/08/2022; Records: 66

- 1 partnerships/ 2429
- 2 public private mix/ 399
- 3 (Public adj3 private adj3 partnership$).mp. 250
- 4 (Public adj3 private adj3 mix$).mp. 492
- 5 (Private adj3 public adj3 collaborat$).mp. 30
- 6 (Private adj3 public adj3 cooperati$).mp. 8
- 7 (Private adj3 public adj3 co-operati$).mp. 11
- 8 (private adj3 public adj3 alliance$).mp. 2
- 9 (private adj3 public adj3 association$).mp. 1
- 10 (private adj3 public adj3 coalition$).mp. 2
- 11 (private adj3 public adj3 union$).mp. 2
- 12 (private adj3 public adj3 consortium$).mp. 0
- 13 (private adj3 public adj3 initiative$).mp. 26
- 14 (private adj3 public adj3 joint$).mp. 6
- 15 (private adj3 public adj3 ownership$).mp. 11
- 16 privatepublic.mp. 1
- 17 publicprivate.mp. 0
- 18 (afghanistan or angola or armenia or armenian or bangladesh or benin or bhutan or bolivia or burkina faso or burkina fasso or upper volta or burundi or urundi or cabo verde or cape verde or cambodia or kampuchea or khmer republic or cameroon or cameron or cameroun or central african republic or ubangi shari or chad or comoros or comoro islands or iles comores or mayotte or democratic republic of the congo or democratic republic congo or congo or zaire or "cote d’ivoire" or "cote d’ ivoire" or cote divoire or cote d ivoire or ivory coast or djibouti or french somaliland or egypt or united arab republic or el salvador or eritrea or eswatini or swaziland or ethiopia or gambia or "georgia (republic)" or georgian or ghana or guam or guatemala or guinea or guinea bissau or haiti or hispaniola or honduras or india or indonesia or timor or jordan or kenya or Kiribati or "democratic people’s republic of korea" or republic of korea or north korea or korea or kosovo or kyrgyzstan or kirghizia or kirgizstan or kyrgyz republic or kirghiz or laos or lao pdr or "lao people's democratic republic" or lesotho or basutoland or liberia or madagascar or malagasy republic or malawi

or nyasaland or mali or micronesia or federated states of micronesia or kiribati or tuvalu or mauritania or moldova or moldovian or mongolia or morocco or ifni or mozambique or portuguese east africa or myanmar or burma or nepal or nicaragua or niger or nigeria or pakistan or papua new guinea or new guinea or philippines or philipines or phillipines or phillippines or rwanda or ruanda or "sao tome and principe" or senegal or sierra leone or melanesia or solomon island or solomon islands or somalia or south sudan or sri lanka or ceylon or sudan or syria or syrian arab republic or tajikistan or tadjikistan or tadzhikistan or tadzhik or tanzania or tanganyika or timor leste or east timor or togo or togolese republic or Tokelau or tunisia or uganda or ukraine or uzbekistan or uzbek or vanuatu or new hebrides or vietnam or viet nam or middle east or west bank or gaza or palestine or yemen or zambia or zimbabwe or northern rhodesia or global south or africa south of the sahara or sub-saharan africa or subsaharan africa or africa, central or central africa or africa, northern or north africa or northern africa or magreb or maghrib or sahara or africa, southern or southern africa or africa, eastern or east africa or eastern africa or africa, western or west africa or western africa or west indies or indian ocean islands or caribbean or central america or latin america or "south and central america" or south america or asia, central or central asia or asia, northern or north asia or northern asia or asia, southeastern or southeastern asia or south eastern asia or southeast asia or south east asia or asia, western or western asia or europe, eastern or east europe or eastern europe or developing country or developing countries or developing nation? or developing population? or developing world or less developed countr* or least developed nation? or least developed population? or least developed world or least developed countr* or less developed nation? or less developed population? or less developed world or lesser developed countr* or lesser developed nation? or lesser developed population? or lesser developed world or under developed countr* or under developed nation? or under developed population? or under developed world or underdeveloped countr* or underdeveloped nation? or underdeveloped population? or underdeveloped world or lower middle income countr* or lower middle income nation? or lower middle income population? or low income countr* or low income nation? or low income population? or lower income countr* or lower income nation? or lower income population? or underserved countr* or underserved nation? or underserved population? or underserved world or under served countr* or under served nation? or under served population? or under served world or deprived countr* or deprived nation? or deprived population? or deprived world or poor countr* or poor nation? or poor population? or poor world or poorer countr* or poorer nation? or poorer population? or poorer world or developing econom* or less developed econom* or lesser developed econom* or under developed econom* or underdeveloped econom* or lower middle income econom* or low income econom* or lower income econom* or low gdp or low gnp or low gross domestic or low gross national or lower gdp or lower gnp or lower gross domestic or lower gross national or lmic or lmics or third world or lami countr* or transitional countr* or emerging economies or emerging nation?).mp. 5470

- 19 or/1-17 2988
- 20 18 and 19 66

### Science Citation Index-Expanded 1900-present, Social Sciences Citation Index 1900-present, Emerging Sources Citation Index 2015-present (Web of Science)

Date Searched: 15/08/2022; Records: 3,061

- #1 TS=(Public NEAR/3 private NEAR/3 partnership$)

- #2 TS=(Public NEAR/3 private NEAR/3 mix$)
- #3 TS=(Private NEAR/3 public NEAR/3 collaborat$)
- #4 TS=(Private NEAR/3 public NEAR/3 cooperati$)
- #5 TS=(Private NEAR/3 public NEAR/3 co-operati$)
- #6 TS=(private NEAR/3 public NEAR/3 alliance$)
- #7 TS=(private NEAR/3 public NEAR/3 association$)
- #8 TS=(private NEAR/3 public NEAR/3 coalition$)
- #9 TS=(private NEAR/3 public NEAR/3 union$)
- #10 TS=(private NEAR/3 public NEAR/3 consortium$)
- #11 TS=(private NEAR/3 public NEAR/3 initiative$)
- #12 TS=(private NEAR/3 public NEAR/3 joint$)
- #13 TS=(private NEAR/3 public NEAR/3 ownership$)
- #14 TS=privatepublic
- #15 TS=publicprivate
- #16 TS=(afghanistan or angola or armenia or armenian or bangladesh or benin or bhutan or bolivia or burkina faso or “burkina fasso” or “upper volta” or burundi or urundi or “cabo verde” or “cape verde” or cambodia or kampuchea or “khmer republic” or cameroon or cameron or cameroun or “central african republic” or “ubangi shari” or chad or comoros or “comoro islands” or “iles comores” or mayotte or “democratic republic of the congo” or “democratic republic congo” or congo or zaire or "cote d’ivoire")
- #17 TS=("cote d’ ivoire" or “cote divoire” or “cote d ivoire” or “ivory coast” or djibouti or “french Somaliland” or egypt or “united arab republic” or “el Salvador” or eritrea or eswatini or swaziland or ethiopia or gambia or "georgia (republic)" or georgian or ghana or guam or guatemala or guinea or “guinea Bissau” or haiti or hispaniola or honduras or india or indonesia or timor or jordan or kenya or Kiribati or "democratic people’s republic of korea" or “republic of korea” or “north korea” or korea or kosovo or Kyrgyzstan)
- #18 TS=(kirghizia or kirgizstan or “kyrgyz republic” or kirghiz or laos or lao pdr or "lao people's democratic republic" or lesotho or basutoland or liberia or madagascar or “malagasy republic” or malawi or nyasaland or mali or micronesia or “federated states of Micronesia” or kiribati or tuvalu or mauritania or moldova or moldovian or mongolia or morocco or ifni or mozambique or “portuguese east Africa” or myanmar or burma or nepal or nicaragua or niger or nigeria or pakistan or papua new guinea)
- #19 TS=( “new guinea” or philippines or philipines or phillipines or phillippines or rwanda or ruanda or "sao tome and principe" or senegal or “sierra leone” or melanesia or “solomon island” or “solomon islands” or somalia or “south sudan” or “sri lanka” or ceylon or sudan or syria or “syrian arab republic” or tajikistan or tadjikistan or tadzhikistan or tadzhik or tanzania or tanganyika or “timor leste” or “east timor” or togo or “togolese republic” or Tokelau or tunisia or uganda or ukraine or uzbekistan or uzbek or Vanuatu)
- #20 TS=( “new Hebrides” or vietnam or “viet nam” or “middle east” or “west bank” or gaza or palestine or yemen or zambia or zimbabwe or “northern Rhodesia” or “global south” or “africa south of the sahara” or “sub-saharan Africa” or “subsaharan Africa” or “africa, central” or “central Africa” or “africa, northern” or “north Africa” or “northern Africa” or magreb or maghrib or sahara or “africa, southern” or “southern Africa” or “africa, eastern” or “east Africa” or “eastern africa” or “africa, western” or “west Africa” or “western Africa”)
- #21 TS=(“west indies” or “indian ocean islands” or caribbean or “central America” or “latin America” or "south and central america" or “south America” or “asia, central” or “central asia” or “asia, northern” or “north asia” or “northern asia” or “asia, southeastern” or “southeastern asia” or “south eastern asia” or “southeast asia” or “south east asia” or “asia,

western” or “western asia” or “europe, eastern” or “east Europe” or “eastern Europe” or “developing country” or “developing countries” or “developing nation?” or “developing population?”)

- #22 TS=(“developing world” or “less developed countr*” or “least developed nation?” or “least developed population?” or “least developed world” or “least developed countr*” or “less developed nation?” or “less developed population?” or “less developed world” or “lesser developed countr*” or “lesser developed nation?” or “lesser developed population?” or “lesser developed world” or “under developed countr*” or “under developed nation?” or “under developed population?” or “under developed world”)
- #23 TS=(“underdeveloped countr*” or “underdeveloped nation?” or “underdeveloped population?” or “underdeveloped world” or “lower middle income countr*” or “lower middle income nation?” or “lower middle income population?” or “low income countr*” or “low income nation?” or “low income population?” or “lower income countr*” or “lower income nation?” or “lower income population?” or “underserved countr*” or “underserved nation?” or “underserved population?” or “underserved world”)
- #24 TS=(“under served countr*” or “under served nation?” or “under served population?” or “under served world” or “deprived countr*” or “deprived nation?” or “deprived population?” or “deprived world” or “poor countr*” or “poor nation?” or “poor population?” or “poor world” or “poorer countr*” or “poorer nation?” or “poorer population?” or “poorer world” or “developing econom*” or “less developed econom*” or “lesser developed econom*” or “under developed econom*” or “underdeveloped econom*”)
- #25 TS=(“lower middle income econom*” or “low income econom*” or “lower income econom*” or “low gdp” or “low gnp” or “low gross domestic” or “low gross national” or “lower gdp” or “lower gnp” or “lower gross domestic” or “lower gross national” or lmic or lmics or “third world” or “lami countr*” or “transitional countr*” or “emerging economies” or “emerging nation?”)
- #1 OR #2 OR #3 OR #4 OR #5 OR #6 OR #7 OR #8 OR #9 OR #10 OR #11 OR #12 OR #13 OR #14 OR #15
- #16 OR #17 OR #18 OR #19 OR #20 OR #21 OR #22 OR #23 OR #24 OR #25
- #26 AND #27

#### CENTRAL, includes EPOC Search Register (The Cochrane Library)

Date Searched: 15/08/2022; Records: 48

- Public-Private Sector Partnerships/
- (Public NEAR/3 private NEAR/3 partnership$) OR (Public NEAR/3 private NEAR/3 mix$) OR (Private NEAR/3 public NEAR/3 collaborat$) OR (Private NEAR/3 public NEAR/3 cooperati$) OR (Private NEAR/3 public NEAR/3 co-operati$) OR (private NEAR/3 public NEAR/3 alliance$) OR (private NEAR/3 public NEAR/3 association$) OR (private NEAR/3 public NEAR/3 coalition$) OR (private NEAR/3 public NEAR/3 union$) OR (private NEAR/3 public NEAR/3 consortium$) OR (private NEAR/3 public NEAR/3 initiative$) OR (private NEAR/3 public NEAR/3 joint$) OR (private NEAR/3 public NEAR/3 ownership$) OR privatepublic OR publicprivate
- (afghanistan or angola or armenia or armenian or bangladesh or benin or bhutan or bolivia or burkina faso or burkina fasso or upper volta or burundi or urundi or cabo verde or cape verde or cambodia or kampuchea or khmer republic or cameroon or cameron or cameroun or central african republic or ubangi shari or chad or comoros or comoro islands or iles comores or mayotte or democratic republic of the congo or democratic republic congo or

congo or zaire or "cote d’ivoire" or "cote d’ ivoire" or cote divoire or cote d ivoire or ivory coast or djibouti or french somaliland or egypt or united arab republic or el salvador or eritrea or eswatini or swaziland or ethiopia or gambia or "georgia (republic)" or georgian or ghana or guam or guatemala or guinea or guinea bissau or haiti or hispaniola or honduras or india or indonesia or timor or jordan or kenya or Kiribati or "democratic people’s republic of korea" or republic of korea or north korea or korea or kosovo or kyrgyzstan or kirghizia or kirgizstan or kyrgyz republic or kirghiz or laos or lao pdr or "lao people's democratic republic" or lesotho or basutoland or liberia or madagascar or malagasy republic or malawi or nyasaland or mali or micronesia or federated states of micronesia or kiribati or tuvalu or mauritania or moldova or moldovian or mongolia or morocco or ifni or mozambique or portuguese east africa or myanmar or burma or nepal or nicaragua or niger or nigeria or pakistan or papua new guinea or new guinea or philippines or philipines or phillipines or phillippines or rwanda or ruanda or "sao tome and principe" or senegal or sierra leone or melanesia or solomon island or solomon islands or somalia or south sudan or sri lanka or ceylon or sudan or syria or syrian arab republic or tajikistan or tadjikistan or tadzhikistan or tadzhik or tanzania or tanganyika or timor leste or east timor or togo or togolese republic or Tokelau or tunisia or uganda or ukraine or uzbekistan or uzbek or vanuatu or new hebrides or vietnam or viet nam or middle east or west bank or gaza or palestine or yemen or zambia or zimbabwe or northern rhodesia or global south or africa south of the sahara or sub-saharan africa or subsaharan africa or africa, central or central africa or africa, northern or north africa or northern africa or magreb or maghrib or sahara or africa, southern or southern africa or africa, eastern or east africa or eastern africa or africa, western or west africa or western africa or west indies or indian ocean islands or caribbean or central america or latin america or "south and central america" or south america or asia, central or central asia or asia, northern or north asia or northern asia or asia, southeastern or southeastern asia or south eastern asia or southeast asia or south east asia or asia, western or western asia or europe, eastern or east europe or eastern europe or developing country or developing countries or developing nation? or developing population? or developing world or less developed countr* or least developed nation? or least developed population? or least developed world or least developed countr* or less developed nation? or less developed population? or less developed world or lesser developed countr* or lesser developed nation? or lesser developed population? or lesser developed world or under developed countr* or under developed nation? or under developed population? or under developed world or underdeveloped countr* or underdeveloped nation? or underdeveloped population? or underdeveloped world or lower middle income countr* or lower middle income nation? or lower middle income population? or low income countr* or low income nation? or low income population? or lower income countr* or lower income nation? or lower income population? or underserved countr* or underserved nation? or underserved population? or underserved world or under served countr* or under served nation? or under served population? or under served world or deprived countr* or deprived nation? or deprived population? or deprived world or poor countr* or poor nation? or poor population? or poor world or poorer countr* or poorer nation? or poorer population? or poorer world or developing econom* or less developed econom* or lesser developed econom* or under developed econom* or underdeveloped econom* or lower middle income econom* or low income econom* or lower income econom* or low gdp or low gnp or low gross domestic or low gross national or lower gdp or lower gnp or lower gross domestic or lower gross national or lmic or lmics or third world or lami countr* or transitional countr* or emerging economies or emerging nation?)

#### Database of disability and inclusion information resources (https://asksource.info/resources/search?)

Date Searched: 15/08/2022; Records: 1

- All fields: public + private + partnership

#### WHOLIS (https://kohahq.searo.who.int/)

Date Searched: 15/08/2022; Records: 0 (when limited to journals)

- All fields: public + private + partnership

#### 3ie International Initiative for Impact Evaluation (https://www.3ieimpact.org/)

Date Searched: 15/08/2022; Records: 721

- Title:(public private) OR Abstract:(public private) OR Keywords:(public private) limited to country filter

### Appendix 3: Eligibility criteria

| **Criteria** | **Include** | **Exclude** |
|---|---|---|
| Population | All urban populations (semi-urban, peri-urban, sub-urban, urban slums) in World Bank defined least developed, low income and lower middle-income countries and territories (https://www.oecd.org/dac/financing-sustainable-development/development-finance-standards/DAC-List-ODA-Recipients-for-reporting-2021-flows.pdf) | All rural populations; also urban populations in upper middle income countries and territories will be excluded – Reason: WRONG POPULATION<br><br>If a study includes both urban and rural populations, BUT data specific to urban populations are not reported separately, it will be excluded – Reason: NO URBAN-SPECIFIC DATA |
| Intervention | Urban health provision in the context of PPPs:<br>Urban health provision – primary/secondary/tertiary healthcare provision, or diagnostic service provision, or provision of primary/secondary/tertiary prevention services in any healthcare or non-healthcare setting.<br>PPP – any long-term contract (i.e. not a one-off event), where the public sector (government and other governmental entities) uses the capacity of the private sector (private companies, cooperatives, charities, non-governmental organisations, and informal private providers) in order to improve and protect the health of populations. | Papers with no PPPs (only one-off events, only public-sector contexts, or only private-sector contexts) will be excluded – Reason: NO INTERVENTION<br><br>PPPs that do not deal with health provision/services/ prevention related care at all will be excluded – Reason: WRONG INTERVENTION |
| Outcome | All outcomes related to Health, Process and WHO System building blocks, as reported | N/A |
| Study design | We will include published journal articles reporting primary research of any of the following types: qualitative and mixed-methods evaluations, cohort study, cross-sectional study, randomised control trial, retrospective study, other intervention study. | Systematic/scoping/narrative reviews will be excluded – Reason: WRONG STUDY DESIGN<br><br>Commentaries, letters, editorials, and conference abstracts will be excluded – Reason: WRONG PUBLICATION TYPE |
| Language | All languages (relying on the team's own language skills/Google translate) | N/A |

### Appendix 4: Synthesis Without Meta-analysis (SWiM) items

| SWiM reporting item | Item description | Page in manuscript where item is reported | Other* |
|---|---|---|---|
| **Methods** | | | |
| 1 Grouping studies for synthesis | 1a) Provide a description of, and rationale for, the groups used in the synthesis (eg, groupings of populations, interventions, outcomes, study design) | Pg. 6 | |
| | 1b) Detail and provide rationale for any changes made subsequent to the protocol in the groups used in the synthesis | Pg. 7 | |
| 2 Describe the standardised metric and transformation methods used | Describe the standardised metric for each outcome. Explain why the metric(s) was chosen and describe any methods used to transform the intervention effects, as reported in the study, to the standardised metric, citing any methodological guidance consulted | Pg. 6 | |
| 3 Describe the synthesis methods | Describe and justify the methods used to synthesise the effects for each outcome when it was not possible to undertake a meta-analysis of effect estimates | Pg. 6 | |
| 4 Criteria used to prioritise results for summary and synthesis | Where applicable, provide the criteria used, with supporting justification, to select the particular studies, or a particular study, for the main synthesis or to draw conclusions from the synthesis (eg, based on study design, risk of bias assessments, directness in relation to the review question) | NA | |
| 5 Investigation of heterogeneity in reported effects | State the method(s) used to examine heterogeneity in reported effects when it was not possible to undertake a meta-analysis of effect estimates and its extensions to investigate heterogeneity | Pgs. 8 and 13 | |
| 6 Certainty of evidence | Describe the methods used to assess the certainty of the synthesis findings | NA | |

| | | |
|---|---|---|
| 7 Data presentation methods | Describe the graphical and tabular methods used to present the effects (eg, tables, forest plots, harvest plots) | Pg. 6 |
| | Specify key study characteristics (eg, study design, risk of bias) used to order the studies, in the text and any tables or graphs, clearly referencing the studies included | |
| **Results** | | |
| 8 Reporting results | For each comparison and outcome, provide a description of the synthesised findings and the certainty of the findings. Describe the result in language that is consistent with the question the synthesis addresses, and indicate which studies contribute to the synthesis | Pgs. 7 – 12 |
| **Discussion** | | |
| 9 Limitations of the synthesis | Report the limitations of the synthesis methods used and/or the groupings used in the synthesis and how these affect the conclusions that can be drawn in relation to the original review question | Pgs. 13 and 14 |

### Appendix 5: Cost calculations and reporting methodology

| | |
|---|---|
| **Costs Currency** | We reported in International Dollars (I$) in 2022 prices to allow comparability across the studies. |
| **Inflated Costs** | We inflated costs using the yearly inflation rates reported by the International Monetary Fund.[1]<br>1. International Monetary Fund. *Inflation rate, end of period consumers prices: Annual percentage change.* 2023. Available:<br>https://www.imf.org/external/datamapper/PCPIEPCH@WEO/OEMDC/ADVEC/WEOWORLD[Accessed 15 August 2023] |

| | |
|---|---|
| **Obtaining Costs** | To obtain costs in I$, inflated costs were divided by the annual purchasing power parity conversion factor reported by the World Bank for each country.[2]<br><br>2. World Bank. *PPP conversion factor, GDP (LCU per international $).* 2023. https://data.worldbank.org/indicator/PA.NUS.PPP [Accessed 15 August 2023] |

### Appendix 6: Characteristics of PPM models in urban settings for tuberculosis care

| Study Year & Author | Country (City) | Intervention provided by PPM model | Public Partner(s) and Role(s) | Private Partner(s) and Role(s) | PPM Funding |
|---|---|---|---|---|---|
| **DIRECT MODELS** | | | | | |
| 2003 Quy (a)<br>2003 Quy (b) | Viet Nam<br>(Ho Chi Minh City) | PPs were provided training, increased supervision, standardised referral and information systems, financial incentives for case detection; participation voluntary, no written agreements, no drugs provided to PPs | NTP: Stewardship; Support; Monitoring; Service | Private health facilities, pharmacies, practitioners: Service | NR |
| 2004 Lonnroth* | Viet Nam<br>(Ho Chi Minh City) | PPs were provided training, increased supervision, standardised referral and information systems, financial incentives for case detection; participation voluntary, no written agreements, no drugs provided to PPs | NTP: Stewardship; Support; Monitoring; Service | Private health facilities, pharmacies, practitioners: Service | External research funds |
| 2006 Krishnan | India (Ballabgarh) | PPs were provided training, standardised referral and information systems, invited to be DOTS providers | RNTCP: Stewardship; Support; Service | Private practitioners: Service | WHO-funded research project |
| 2008 Lagrada | Philippines (NR) | Private health facilities certified as DOTS centres | DOH: Stewardship | Private health facilities: Service | NR |
| 2009 Ahmed | Pakistan (Thatta) | PPs were provided training, financial incentives to refer people for case detection | NTP (via local offices): Stewardship; Support; Monitoring; Financing | Private health facilities: Service | NTP and WHO |
| 2009 Pantoja (a)<br>2009 Pantoja (b) | India (Bengaluru) | Pre-PPM (1997-2001), DOTS mainly in public facilities; PPM phase I (2001-2003), DOTS expanded to public & private medical colleges, NGOs, other public facilities; PPM phase II (post-2003), DOTS scaled up to PPs, NGOs, more facilities | RNTCP: Stewardship; Support; Monitoring; Service | Private health facilities, practitioners, NGOs: Service | NR |
| 2010 Quelapio | Philippines (Manila) | People on MDR-TB treatment decentralised to DOTS facilities run by FBOs, NGOs and public health centres referred to as treatment sites | NTP: Stewardship; Support; Monitoring; Financing; Service | Private health facilities, practitioners, NGOs: Service | NTP with support from Global Fund, Eli Lilly, WHO, CDC |

| | | | | | |
|---|---|---|---|---|---|
| 2011 Lal | India<br>(14 major cities) | PPs were provided training, increased supervision, standardised referral and information systems, quality-assured free drugs; no incentives provided | RNTCP: Stewardship; Support; Service | Private health facilities, practitioners, NGOs: Service | NR |
| 2011 Pradhan | India<br>(Pimpri Chichwad) | PPs were provided training, increased supervision through HVs, standardised referral and information systems, quality-assured free drugs | RNTCP: Stewardship; Support; Monitoring; Financing; Service | Private health facilities, practitioners, NGOs: Service | RNTCP |
| 2013 Daboer | Nigeria<br>(Plateau state) | Engagement of all care providers through PPM; no further information | NTP: Unclear | Private health facilities: Unclear | NR |
| 2013 Daniel | Nigeria (Lagos) | PPs were provided training, increased supervision, standardised referral and information systems, quality-assured free drugs, and diagnostic supplies | LSTBLCP: Support, Monitoring | Private health facilities: Service | NR |
| 2014 Kielmann | India<br>(Sunder Nagar⁺) | PPs were provided training, increased supervision through HVs, standardised referral and information systems; served as DOTS providers | RNTCP: Stewardship; Support; Monitoring; Service | Private practitioners: Service | RNTCP |
| 2014 Subramaniyam | India<br>(Bengaluru) | PPs were provided increased supervision, standardised referral and information systems, quality-assured free drugs for people diagnosed with TB | RNTCP: Stewardship, Service | Private health facility: Monitoring; Service | NR |
| 2016 Salve | India (NR) | PPs were provided increased supervision, standardised referral and information systems, quality-assured free drugs; served as DOTS providers with financial incentives | RNTCP: Stewardship; Support; Monitoring; Service | Private practitioners: Monitoring, Service | NR |
| 2017 Khan | Pakistan (Lahore) | PPs were provided training, increased supervision, standardised referral and information systems, quality-assured free drugs | NTP: Stewardship; Support; Service | Private health facilities, practitioners, and NGOs: Service | NR |
| 2017 Qader | Afghanistan (Kabul) | The urban DOTS model in Kabul focuses on four major intervention areas: (1) building the capacity of the NTP and PPs; (2) expanding DOTS in public and private facilities; (3) improving management and drug supply at facilities; and (4) improving surveillance, supervision, and monitoring | NTP: Stewardship; Support; Monitoring; Service | Private health facilities: Service | NTP with support from USAID, WHO & private sector |

| | | | | | |
|---|---|---|---|---|---|
| 2017 Reviono | Indonesia (Central Java) | Hospital DOTS Linkage included training, increased supervision, standardised referral and information systems, quality-assured free drugs, DOTS centres | NTP: Monitoring | Private health facilities: Service | NR |
| 2018 Awan | Pakistan (Karachi) | The social business model utilised community-based screeners at 180 private clinics (both formal and informal) to screen and refer people for TB testing to three purpose-built TB centres called 'Sehatmand zindagi' ('healthy life') | PTP: Support; Monitoring; Service | Private health facilities: Service | NR |
| 2018 Yellappa | India (Tumkur city) | PPs were provided training, increased supervision, standardised referral and information systems, TB IEC | RNTCP: Stewardship; Support; Service | Private practitioners: Service | NR |
| 2019 Hadisoemarto<br>2021 Hadisoemarto | Indonesia (Bandung) | The intervention will comprise: (1) An electronic referral and notification system; (2) Education about signs and symptoms of TB management; (3) An individualised plan for diagnostic and management pathways | NTP: Support; Monitoring; Service | Private practitioners: Service | Research funds from HRC-NZ & Indonesian Ministry |
| 2019 Hemavarneshwari | India (Bengaluru) | PPs were provided training by a MSW on case finding and referral; poster and case referral book developed for PPs | RNTCP: Stewardship; Support | Private practitioners (with training from a private hospital): Service | State Tuberculosis Office, Bengaluru |
| 2020 Paul | Bangladesh (Dhaka) | Persons attending PPM TB screening centres in urban Dhaka were also offered free blood glucose testing | NTP: Support; Service | Private practitioners: Service | NR |
| 2020 Thu | Viet Nam (Haiphong province) | PPs were provided training, standardised referral and information systems, financial incentives for case detection and treatment success | TB Control Programme: Stewardship; Support; Monitoring; Service | Private health facility: Monitoring; Service | TB REACH-funded Zero TB Vietnam initiative |
| 2021 Oladimeji | Nigeria (Lagos) | Facilities with MoU were provided training, increased supervision, standardised referral and information systems, quality-assured free drugs and supplies | LSTBLCP: Stewardship; Support; Monitoring; Financing; Service | Private health facilities: Service | NR |
| **INTERFACE MODELS** | | | | | |
| 2001 Murthy | India (Chennai) | PPs were provided training, standardised referral and information systems; served as DOTS providers | RNTCP: Stewardship; Support; Service | Private health facilities, practitioners (referral model via a private | WHO & DFID |

| | | | | | |
|---|---|---|---|---|---|
| | | | | hospital): Stewardship; Support; Monitoring; Service | |
| 2002 Hurtig | Nepal (Kathmandu) | Service linkage project – PPs were provided training, standardised referral and information systems; no drugs provided to PPs | NTP: Stewardship; Support; Service | Private pharmacies, practitioners via a research organisation: Service | Research funds from DFID |
| 2003 Rangan | India (Mumbai) | PPs were provided training, TB IEC; decentralization of DOTS and support from External Quality Assurance Scheme | RNTCP: Stewardship; Service | Private health facilities, practitioners via NGO: Support; Monitoring; Service | NR |
| 2003 Tupasi<br>2006 Tupasi | Philippines (Manila) | DOTS-Plus for MDR-TB (individualized DOTS regimen based on drug susceptibility testing) implemented at a private health facility | NTP: Stewardship; Monitoring; Service | Private health facility (with support from a research organisation): Support; Service | Philippine Charity Sweepstakes & UNDP/WHO/TDR Programme |
| 2004 Arora | India (New Delhi) | PPs were provided training, increased supervision, standardised referral and information systems, quality-assured free drugs | RNTCP: Stewardship; Support; Monitoring | Private practitioners via Medical Association: Support; Service | NR |
| 2004 Lonnroth* | India (New Delhi) | PPs were provided training, increased supervision, standardised referral and information systems, quality-assured free drugs | NTPs: Stewardship; Support; Monitoring; Financing; Service | Private practitioners via Medical Association: Service | Ministry of Health |
| | Kenya (Nairobi) | PPs were provided training, increased supervision, standardised referral and information systems, drugs supplied by a donor agency | NTP: Stewardship; Support; Monitoring; Service | Private practitioners via NGO: Support; Service | Donor agency |
| 2004 Newell<br>2005 Newell | Nepal (Lalitpur municipality) | PPs were provided training, increased supervision, standardised referral and information systems, quality-assured free drugs; DOTS and tracing of patients who missed appointments by NGOs | NTP: Stewardship; Support; Monitoring; Financing | Private health facilities, practitioners, NGOs (with support from a research organisation): Support; Monitoring; Financing; Service | Research funds from DFID |
| 2005 Ambe | India (Mumbai) | PPs were provided training, increased supervision, standardised referral and information systems, quality-assured free drugs; served as DOTS providers | RNTCP: Stewardship; Support; Monitoring; Financing; Service | Private health facilities, practitioners, NGOs via Medical Association, and | RNTCP with support from WHO |

| | | | | | |
|---|---|---|---|---|---|
| | | | | core NGOs: Support; Service | |
| 2005 Lambert | Bolivia (Cochabamba) | Collaboration phase 1: recommendation to member pharmacies to stop selling TB drugs; phase 2: refer clients seeking TB drugs to public services | NTP: Stewardship; Support; Service | Private pharmacies via Pharmacists Association: Service | NR |
| 2006 Floyd | India (Hyderabad) | PPs were provided training, increased supervision through private hospital staff, standardised referral and information systems, quality-assured free drugs | RNTCP: Support; Financing | Private practitioners (referral model via private hospital): Support; Service | RNTCP |
| | India (New Delhi) | PPs were provided training, increased supervision, standardised referral and information systems, quality-assured free drugs | RNTCP: Support; Financing | Private practitioners via Medical Association: Service | RNTCP |
| 2006 Maung | Myanmar (Kyaukse township) | PPs were provided training, increased supervision, standardised referral and information systems; served as DOTS providers (no incentives) | NTP: Stewardship; Support; Monitoring; Service | Private practitioners (with support from research organisation): Service | NTP with partial support from Global Fund |
| 2007 Irawati | Indonesia (Yogyakarta) | Hospital DOTS linkage pilot provided training, increased supervision, standardised referral and information systems, quality-assured free drugs, DOTS centres | NTP: Stewardship; Support; Monitoring; Service | Private health facilities via an Association: Service | University of Alabama, USAID, KNCV Foundation & TBCTA |
| 2007 Karki | Nepal (Kathmandu) | PPs were provided standardised referral and information systems; served as DOTS providers; private facilities invited to become DOTS centres with MoU | NTP: Stewardship; Support; Monitoring; Service | Private practitioners via private health facilities and NGOs: Support; Service | NR |
| 2008 Chakaya | Kenya (Nairobi) | PPs were provided training for TB and HIV, increased supervision, standardised referral and information systems, drugs obtained by patients using a prepayment scheme or free from the state | NTP: Stewardship; Support; Monitoring; Service | Private practitioners via NGO: Support; Service | CDC, CHF International & Sanofi-Aventis |
| 2009 Krishnan | India (Chennai) | PPs were provided training by NGO, increased supervision, standardised referral and information systems, quality-assured free drugs | RNTCP: Stewardship; Support; Service | Private practitioners via NGO (with support from a research organisation): Support; Service | Global Fund |

| | | | | | |
|---|---|---|---|---|---|
| 2012 Bell<br>2015 Bell | Cambodia (Phnom Penh) | The programme promoted pharmacy-initiated assessment and referral of people with TB symptoms to public-sector DOTS centres providing free diagnosis and treatment; pharmacies elected to join the referral programme | CENAT: Stewardship; Support; Monitoring | Pharmacists Association and private pharmacies via NGOs: Service | Global agencies |
| 2012 Khan | Pakistan (Karachi) | A year-long communications campaign by private hospital; screening at hospital by community laypeople with financial incentives for case detection | NTP: Stewardship; Support | Private health facilities via a private hospital with community screeners: Support; Service | TB REACH-funded Stop TB partnership |
| 2012 Naqvi | Pakistan (Karachi) | Mapping PPs and cluster formations, contracting laboratory networks, training in DOTS, increased supervision, quality-assured free drugs | NTP: Stewardship; Service | Private practitioners via a research university: Support; Monitoring | NR |
| 2012 Zafar Ullah (a) | Bangladesh (Dhaka) | PPs were provided training by NGO, increased supervision, standardised referral and information systems; public DOTS centres strengthened | NTP: Service | Private practitioners via NGOs: Stewardship; Support; Monitoring; Service | Research funds from DFID |
| 2012 Zafar Ullah (b) | Bangladesh (Dhaka) | Workplace DOTS services by NTP/NGOs and garment association | NTP: Stewardship; Support; Monitoring; Service | Private health facilities managed by industry (via support from NGOs): Support; Service | Research funds from DFID |
| 2014 Engel | India (Hyderabad) | PPs were provided training, increased supervision through private hospital staff, standardised referral and information systems, quality-assured free drugs | Unclear: Not clearly reported | Private practitioners (referral model via private hospital): Support; Service | NR |
| | India (Mumbai) | NGO with a civil society mandate acted as an intermediary between private practitioners and the TB programme | RNTCP: Stewardship; Monitoring; Service | Private practitioners via NGO: Support; Service | NR |
| | India (Kerala state) | Sensitisation of PPs by Indian Medical Association; no further information | RNTCP: Stewardship; Monitoring; Service | Private practitioners via Medical Association: Service | NR |
| 2015 Pethani | Pakistan (Karachi) | PPs were provided training, increased supervision, standardised referral and information systems, quality-assured free drugs; served as DOTS providers | NTP: Stewardship; Support; Monitoring; Service | Private practitioners via a research university: Stewardship; Support | WHO |

| | | | | | |
|---|---|---|---|---|---|
| 2015 Ramaiah | India (Kanchipuram) | NGO as the sole service provider of RNTCP services in a defined geographic area; increased supervision of PPs | RNTCP: Stewardship; Support; Monitoring; Financing | Private practitioners via NGO: Support; Service | RNTCP |
| 2016 Bronner Murrison | India (Chennai) | PPs refer people with TB to NGO centres to receive free DOTS under the supervision of NGO staff | RNTCP: Service | Private practitioners via NGO: Support; Service | NR |
| 2017 Lestari | Indonesia (Bandung City) | PPs were provided training and mobile phone application by research team, increased supervision | NTP: Stewardship; Support; Monitoring | Private practitioners via a research university: Support; Service | Research funds from University of Otago, NZ |
| 2018 Chadha | India (Bengaluru) | PPs were provided training by TB Unit, increased supervision, standardised referral and information systems | RNTCP: Stewardship; Support; Monitoring | Private health facilities via Medical Association: Service | NR |
| 2018 McDowell | India (4 major cities) | Hub and spoke model, linking laboratories with public and private sector providers for free-of-charge Xpert testing for paediatric presumptive TB | RNTCP: Stewardship; Service | Private health facilities and practitioners (via a non-profit organisation): Support; Service | USAID |
| 2019 Ananthakrishnan | India (Chennai) | PPs were provided training, increased supervision by NGO, standardised referral and information systems; choice for PP/ patient to decide where patient will receive treatment | RNTCP: Stewardship; Support; Monitoring; Service | Private practitioners via NGO: Support; Service | External funding from USAID & Stop TB Partnership |
| 2019 Arinaminpathy<br>2021 Arinaminpathy | India (Mumbai, Patna) | PPs were provided training by NGO, increased supervision, standardised referral and information systems, quality-assured free drugs, financial incentives | RNTCP: Stewardship; Support; Monitoring; Financing; Service | Private practitioners via PPIA NGOs: Support; Service | Government with World Bank & Global Fund |
| 2019 Daftary | India (Patna) | (1) Pharmacy training, (2) TB screening and e-referrals for testing and consultation, (3) Incentives for referral completion and TB diagnosis, (4) Reminders, and (5) Field support, supervision, and monitoring by PPIA staff | RNTCP: Stewardship; Support; Financing | Private pharmacies and practitioners via PPIA: Support; Service | NR |
| 2020 Ramasamy (a)<br>2020 Ramasamy (b) | India (Bengaluru) | Liaison model delivering educational intervention to member pharmacies support the delivery of DOTS | RNTCP: Stewardship; Support; Monitoring; Service | Private pharmacies via an Association: Support; Service | NR |

| | | | | | |
|---|---|---|---|---|---|
| 2020 Shibu | India (Mumbai) | PPIA service delivery model: field officers acted as marketing and relationship managers for PPs to ensure optimal uptake of PPIA services; community NGOs engaged to support sputum transportation, monitor treatment adherence, and facilitate reporting | RNTCP: Stewardship; Service | Private pharmacies & practitioners via PPIA: Support; Financing; Service | Gates Foundation |

* 2004 Lonnroth reports 3 PPM models – 1 direct and 2 with interface; + Pseudonym

Abbreviations (ordered alphabetically): CDC – Centres for Disease Control and Prevention; DFID – Department for International Development; DOH - Department of Health; DOTS – Directly Observed Therapy Short-course; FBO – Faith-Based Organisation; HIV – Human Immunodeficiency Virus; HV – Health Visitor; HRC-NZ – Health Research Council of New Zealand; IEC – Information, Education and Communication; LSTBLCP – Lagos State Tuberculosis, Buruli Ulcer, and Leprosy Control Program; MDR-TB – Multidrug-Resistant Tuberculosis; MoU – Memorandum of Understanding; MSW – Medical Social Worker; NGO – Non-Governmental Organisation; NR – Not Reported; NTP – National Tuberculosis Programme; PP – Private Provider; PPIA – Public-Private Interface Agency; PPM – Public-Private Partnership; PTP – Provincial Tuberculosis Programme; RNTCP – Revised National Tuberculosis Control Programme; TB – Tuberculosis; TBCTA – Tuberculosis Coalition for Technical Assistance; TDR – Special Programme for Research and Training in Tropical Diseases; UNDP – United Nations Development Programme; USAID – The United States Agency for International Development; WHO – World Health Organization.

### Appendix 7: Forest plot (without meta-analysis) of studies reporting TB treatment success comparing PPM DOTS with public DOTS/private non-DOTS

- 10 studies compare TB treatment outcomes of PPM DOTS with public DOTS/private non-DOTS, but CI not reported/could not be calculated in five studies; only studies with CIs have been plotted in the figure below:

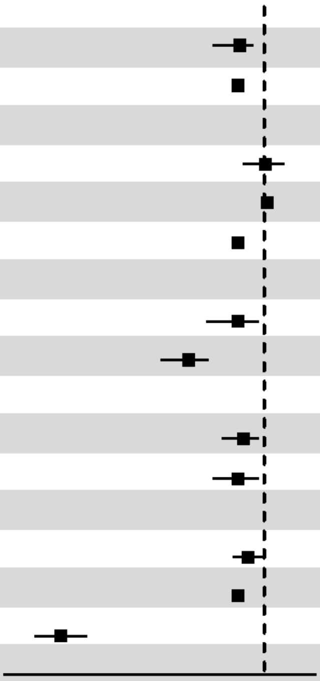

| Studies with comparison | TS (%) | LB | UB |
|---|---|---|---|
| **2005 Ambe (India)** | | | |
| PPM DOTS | 85.4 | 80.2 | 88.1 |
| Public DOTS | 85 | 84.3 | 86.1 |
| **2006 Maung (Myanmar)** | | | |
| PPM DOTS | 90.4 | 83.1 | 94.8 |
| Public DOTS, Intervention | 90.6 | 84.5 | 94.6 |
| Public DOTS, Control | 85.1 | 79.9 | 89.2 |
| **2008 Chakaya (Kenya)** | | | |
| PPM DOTS | 85 | 80.6 | 88.5 |
| Public DOTS | 75.4 | 74.3 | 76.4 |
| **2011 Pradhan (India)** | | | |
| PPM DOTS | 86 | 82 | 89 |
| Public DOTS | 85 | 80 | 89 |
| **2015 Ramaiah (India)** | | | |
| PPM DOTS | 87 | 84 | 90 |
| Public DOTS | 85 | 84 | 85 |
| Private non-DOTS | 51 | 46 | 56 |

### Appendix 8: Cost and cost-effectiveness studies (costs reported as I$, 2022 prices)

#### Cost studies

| Ref | Setting | Intervention | PPP characteristics | Methodological approach | Cost |
|---|---|---|---|---|---|
| 2001 Murthy | India, Hyderabad | Public-private mix model of DOTS | Goal: DOTS implementation<br>Roles<br>Public - The Revised National TB Control Programme (RNTCP): provision of medicines, training and laboratory supplies<br>Private - Private practitioners PPs), charitable trust hospital (Mahavir Hospital): Referral for TB test (PPs), TB diagnosis and DOTs provision, case recording and reporting (Mahavir Hospital) | Costs<br>Patient costs: out-of-pocket expenditure on fees, transport, diagnostic investigations, medications and indirect costs (income lost)<br><br>Outcome<br>Average of TB treatment | Average treatment cost (out of pocket, income lost)<br>Mahavir Hospital (PPP) vs government RNTCP centre (non-PPP)<br>Pre-TB Diagnosis: 37.80 vs 151.19<br>Treatment: 7.56 vs 83.16<br>Income lost: 1.4 vs 2.8 months' lost wages |
| 2007 Karki | Nepal, Lalitpur | PPP scheme | Goal: Increase TB treatment quality and coverage<br>Roles<br>Public - National TB Programme (NTP) and District Public Health Office (DPHO): provision of training for laboratory staff and DOTs supervisors, supervision of workers involved in TB control, provision and distribution of medicines, and monitoring Treatment Centres (TCs) regarding the adherence to the NTP guidelines<br>Private - Semi-governmental hospital, NGOs, Nepal Anti-TB Association (NATA), private nursing home: refer patients to the DOTS centres | Costs<br>Provider costs (DPHO and TCs): Start-up (managerial set-up of the scheme, training of private practitioners, TC staff and volunteers) and recurrent (costs of staff, procurement and delivery of medicines, refresher training courses and regular DOTS workshop<br>Volunteers: start-up (time spent on training) and recurrent (travel expenses and opportunity costs of time spent implementing activities)<br>Patient costs: travel and time lost<br><br>Outcome<br>Costs of implementing PPP TB, average cost of TB treatment | Total cost per patient of implementing PPP TB<br>Provider<br>Start-up costs: 200.99<br>Recurrent costs: 337.31<br>Total costs: 538.30<br>Volunteer<br>Start-up costs: 4.37<br>Recurrent costs: 12.23<br>Total costs: 16.60<br><br>Total cost of implementing PPP TB<br>Provider<br>Start-up costs: 103,827,54<br>Recurrent: 195,396.87<br>Total: 299,224.41<br>Volunteer<br>Start-up: 2,341.89<br>Recurrent: 4,496.15<br>Total: 6,838.04<br><br>Average treatment cost (out of pocket and time lost): 1,588.69 |
| 2009 Pantoja | India, Bangalore | PP mix for TB care and control | Goal: Ensure good quality diagnosis and treatment at low cost across the entire health system, including all public as well as private health providers | Costs<br>Patient costs: consultation fees, medicines, diagnostic tests, hospitalisation, | Average treatment cost (out of pocket and time lost)<br>Average cost before treatment in the RNTCP: 897.80 |

| Ref | Setting | Intervention | PPP characteristics | Methodological approach | Cost |
|---|---|---|---|---|---|
| | | | Roles<br>Public - The Revised National TB Control Programme (RNTCP): Supply and/or subsidise the cost of medicines and diagnostics for patients who are treated by private as well as public sector providers<br>Private - Private practitioners, non-governmental organisations and the corporate sector: Referral to RNTCP facilities for diagnosis and treatment | transportation, days lost from work due to health seeking<br><br>Outcome<br>Average cost incurred by patients before treatment in the RNTCP and outside RNTCP; costs as a proportion of annual household income per capita | Average cost of treatment within RNTCP: 130.03<br>Average cost of treatment outside RNTCP: 786.35<br><br>Costs as a proportion of annual household income per capita<br>Low and middle standard of living households: 53%<br>Other households: 41% |
| 2017 Lestari | Indonesia, Bandung | PPP for TB case detection | Goal: Collaboration with private practitioners for TB detection, diagnosis and treatment<br>Roles<br>Public - National TB control programme: Develop referral and report-back system using a mobile phone application; provide training to private practitioners on case management, report and recording and mobile phone application; monitoring<br>Private - Private practitioners: Provide treatment, and implement the mobile phone application | Costs<br>Health system costs: recurrent and capital costs<br><br>Outcomes<br>Total costs of PPP implementation and maintenance, cost per person with TB symptoms, cost per TB cases diagnosed, cost per private practitioner | Total costs<br>Start-up costs: 9,258.75<br>Recurrent costs: 4,968.45<br>Total costs: 14,227.21<br>Maintenance costs (1-year): 1,529.85<br>Cost per person with TB symptoms: 395.20<br>Cost per TB cases diagnosed: 836.91<br>Cost per private practitioner: 1,185.60 |
| 2019 Daftary | India, Patna | Community pharmacists to improve TB case finding | Goal: improving pharmacies' engagement in TB screening and referral<br>Roles<br>Public - Provision of interactive training workshops on TB symptomology, screening and diagnostic testing, antibiotic stewardship; study referral practices and documentation; E-health messages to reinforce training; supervision and monitoring; provision of incentives to the pharmacists (completed chest X-Ray referral, case notification)<br>Private - Pharmacists: TB screening and e-referral | Costs<br>Health system costs: incentives for chest X-Ray , referrals and case notifications; e-health text messaging services and human resources<br>Outcomes<br>Total cost of the intervention, cost per case detected | Total cost: 123,535.95<br>Cost per case detected: 424.42 |

### Cost-effectiveness studies

| Ref | Location | Interventions | PPP characteristics | Methodological approach | Cost-effectiveness |
|---|---|---|---|---|---|
| 2006 Floyd | India, New Delhi and Hyderabad | • Public–Private Mix DOTS (PPM-DOTS)<br>• Public sector DOTS<br>• Non-DOTS treatment in the private sector (private non-DOTS) | <u>Goal:</u> Implementation of DOTS in the private sector<br><u>Roles</u><br>Public - Revised National Tuberculosis Control Programme (RNTC): Provision of a budget (start-up and routine implementation activities), supply of free drugs and laboratory supplies<br>Private - Not-for-profit institution and Delhi Medical Association: Managing PPM-DOTS implementation, reporting cases detected and treatment outcomes | <u>Perspective</u><br>Public sector, patients and attendants and private practitioners<br><u>Outcome</u><br>Incremental cost per patients successfully treated<br><u>Model</u><br>Multivariate uncertainty analysis | ICER per patients successfully treated<br><br>PM-DOTS vs public sector DOTS<br>Hyderabad<br>Public sector perspective: cost-saving<br>Provider perspective: 3 (CI not available)<br>Societal perspective: 56 (95% CI: 1–110)<br><br>PPM-DOTS vs private sector non-DOTS<br>Delhi<br>Public sector perspective: 95 (95% CI: 85–107)<br>Provider perspective: 211 (95% CI: 189–236)<br>Hyderabad<br>Public sector perspective: 56 (95% CI: 51–61)<br>Provider perspective: 123 (95% CI: 112–135)<br>Societal perspective: 0 (95% CI: -45 to 45) |
| 2015 Ramaiah | India, Tamil Nadu | • PPM-DOTS<br>• Public sector DOTS<br>• Private sector non-DOTS | <u>Goal</u>: Implementation of DOTS in the private sector<br><u>Roles</u><br>Public - RNTC: Provision of budget ,drugs, laboratory supplies, training and DOT provider incentives, TB diagnosis and DOTS provision<br>Private - Not-for-profit institution: TB diagnosis and DOTS provision | <u>Perspective</u><br>Public provider and societal<br><u>Outcome</u> Incremental cost per patients successfully treated<br><u>Model</u><br>Decision tree | ICER per patients successfully treated<br><br>PPM-DOTS vs public sector DOTS<br>Provider perspective: –21 (95% CI:–44 to –3)<br>Societal perspective: -278 (95% CI:-312-256<br><br>PPM-DOTS vs private sector non-DOTS<br>Provider perspective: 226 (95% CI: 184-292),<br>Societal perspective: cost-saving |
| 2006 Tupassi | Philippines | DOTS-Plus Pilot Project | <u>Goal</u>: improve multidrug resistant TB diagnosis (i.e., culture and drug susceptibility testing) and treatment (second- as well as first-line drugs)<br><u>Roles</u><br>Public - Department of Health and the local government: diagnosis and referral of chronic MDR-TB patients<br>Private - Makati Medical Centre: Provision of DOTs and follow-up of defaulted patients | <u>Perspective</u><br>Societal<br><u>Outcomes:</u> total DALYs lost for each strategy, the total DALYs gained by DOTS-Plus, and the cost per DALY gained by DOTS-Plus<br><u>Model</u>: Transmission model, multivariate uncertainty analysis, Monte Carlo simulation | DOTS-Plus project available vs project not-available<br>Total DALYs gained: 2,773 (95% CI: 1,247 - 6,385)<br>Cost per DALY gained<br>Health system perspective: 179 (95% CI:46 - 334)<br>Societal perspective: 242 (95% CI: 85- 426) |

| | | | | | |
|---|---|---|---|---|---|
| 2009 Pantoja | India, Bangalore | ● public-private mix (PPM) for TB care and control Phase I [1] and II [2] implemented<br>● PPM Phase I implemented<br>● Pre-PPM [3] | <u>Goal</u>: Ensure good quality diagnosis and treatment at low cost across the entire health system, including all public as well as private health providers<br><u>Roles</u><br>Public<br>The Revised National TB Control Programme (RNTCP): Supply and/or subsidise the cost of medicines and diagnostics for patients who are treated by private as well as public sector providers<br>Private<br>Private practitioners, non-governmental organisations and the corporate sector: Referral to RNTCP facilities for diagnosis and treatment | <u>Perspective</u><br>Provider and societal<br><u>Outcome</u><br>Incremental cost per patients successfully treated<br><u>Model</u><br>Multivariate uncertainty analysis | ICER per patients successfully treated<br>PPM Phase I and II implemented vs PPM Phase I implemented<br>Provider perspective: 93 (95% CI: 89–102)<br>Societal perspective: cost-saving<br>PPM Phase I and II implemented vs Pre-PPM1<br>Provider perspective: 69 (95% CI: 66–70)<br>Societal perspective: cost-saving |
| 2021 Arinaminpathy | India, Mumbai and Patna | 'Public–Private Interface Agency' (PPIA):<br>Linear increase in provider recruitment from 2014 to 2017 vs pre-2014 levels<br><br>Different coverages (25%, 50%, 75%)<br><br>Different services provided (diagnosis or treatment) | <u>Goal</u><br>Provide high-quality diagnostic tests in the private sector, offer free TB drugs, and adherence support mechanisms to TB patients to maximise treatment completion and facilitates TB notification<br><u>Roles</u><br>Public<br>India's Central TB Division, India National Tuberculosis Elimination Programme: Provision of training to improve TB diagnosis, and incentives for symptomatic tested, TB patients diagnosed and TB patients completing treatment<br>Private<br>Non-Governmental Organisations: Provision of free TB treatment and link to a call centre for adherence monitoring and support | <u>Perspective</u><br>Provider<br><u>Outcome</u><br>Incremental cost per disability adjusted life years (DALY) averted<br><u>Model</u><br>Transmission modelling (WHO-CHOICE [4] and country-specific Thresholds [5] including opportunity costs) | ICER per DALYs averted<br>All coverages<br>Mumbai: PPIA is cost-effective in all thresholds<br>Patna: Cost-effective for the WHO-CHOICE; not cost-effective for the country-specific threshold<br>PPIA focuses on adherence support<br>Mumbai: 30.5 (95% CI: 3.46–79.7), best value, cost-effective in all thresholds<br>Patna: 72.6 (95% CI: 29.6–157), best value, cost-effective in all thresholds<br>PPIA focuses on quality of TB diagnosis<br>Mumbai: 441 (95%CI: 319–601)<br>Patna: 803 (95% CI: 566–1120)<br>PPIA focuses on adherence and TB diagnosis<br>Mumbai: 228 (95%CI: 159–320)<br>Patna: 564 (95% CI: 409–775) |

ICER= incremental cost-effectiveness ratio

[1] PPM Phase I: RNTCP started to engage with public and private medical colleges, and increased collaboration with NGOs as well as public sector entities not previously involved in the RNTCP; [2] PPM Phase II: scale up and intensify PPM in 14 large cities; [3] Pre-PPM phase: DOTS implemented almost exclusively in the facilities of the Ministry of Health; [4] WHO-CHOICE Cost-effectiveness threshold, highly cost-effective: Mumbai $1,500, Patna $530; WHO-CHOICE, cost-effective: Mumbai $4,500, Patna $1,590; [5] Country-specific threshold, Mumbai and Patna $290
